# Understanding repair and replacement of prosthetic limbs using routinely-collected data: a retrospective study over three decades in Cambodia

**DOI:** 10.1101/2024.10.15.24315396

**Authors:** A S Dickinson, L Gates, C D Metcalf, C Owen, S Kheng, T Heang, B Sam, C Harte, S Simpson, P R Worsley, C Ostler, M Donovan-Hall, A A Channon

**Author notes:** Corresponding Author. 7/5004/MP7, Mechanical Engineering Department, Faculty of Engineering & Physical Sciences, University of Southampton, UK.

## Abstract

Prosthetic limbs deliver major quality of life and socioeconomic benefits for people with amputation, particularly in low-resource settings. The value of administrative data analysis to enable sustainable health care improvement is established, but there has been limited research into the failure, repair, and replacement of prosthetic limbs. Survivorship data is sparse and highly variable, and rarely addresses differences between demographic groups.

Therefore, we investigated the distribution of time between device delivery, repair and replacement for a Cambodian cohort, considering the influence of a range of service delivery, user demographic and health characteristics. We conducted Kaplan-Meier survival analysis, and a Cox model to compare repair and replacement likelihood between groups.

The study explored 14,822 device deliveries to 6,986 clients, with median 3 devices/person (interdecile range IDR 1–9), and 22,205 repairs, median 1 repair/device (IDR 0–4). The median device survival before repair was 237 days (IDR 38–854), and replacement was 727 days (IDR 208–2154). Devices used by people in more active occupations were repaired earlier, and devices were replaced earlier when used by children, replaced later for upper- than lower-limb devices, and replaced earlier for volume change than for wear and tear. Several less intuitive trends were revealed such as different preferences or capacities between clinics for device repair vs. replacement, and earlier device repair and replacement for women than men.

Prosthetic limb repair and replacement is influenced both by the device’s durability and the user’s access to well-resourced physical rehabilitation services. A worn-out device may indicate poor quality, or the opposite: that it fitted well and enabled great mobility. However, such analysis may enable us to identify groups who are less well-served by current devices or rehabilitation models, and contribute to cost effectiveness analysis of current services. Furthermore, the findings represent benchmark data against which engineers could measure new technologies, to ensure that innovation justifies its inherent risk by offering a genuine improvement which balances functionality, cost, and durability.

## Introduction

Great social and economic benefits are offered by prosthetics and physical rehabilitation services, both to the individual and to society. However, major negative emotional and socioeconomic impacts can arise for a person with amputation when their prosthetic limb ceases to fit, is damaged, requires maintenance, or fails [1–4]. Prosthetic limb provision is inevitably iterative, where the amputated limb matures and stabilises following surgery or congenital changes. Indeed, clinic or workshop visits during early rehabilitation are most associated with socket suspension due to discomfort, volume, and shape changes. Studies across a range of amputation levels in two high-income settings report an average of 9 visits to prosthetics services per year in the early phases of prosthetic fitting [5,6]. Replacements due to loss of socket fit may continue at lower frequency throughout the person’s life, and clinic visits are also required for repair and replacement of the other prosthetic components [6]. Under repetitive loading, failures may occur in the prosthetic componentry whose certification requires a fatigue life over 3 million cycles [7]. Similarly, modular adapter interfaces between components may become loose, need adjustment, or replacement (Figure 1).

**Figure 1:**
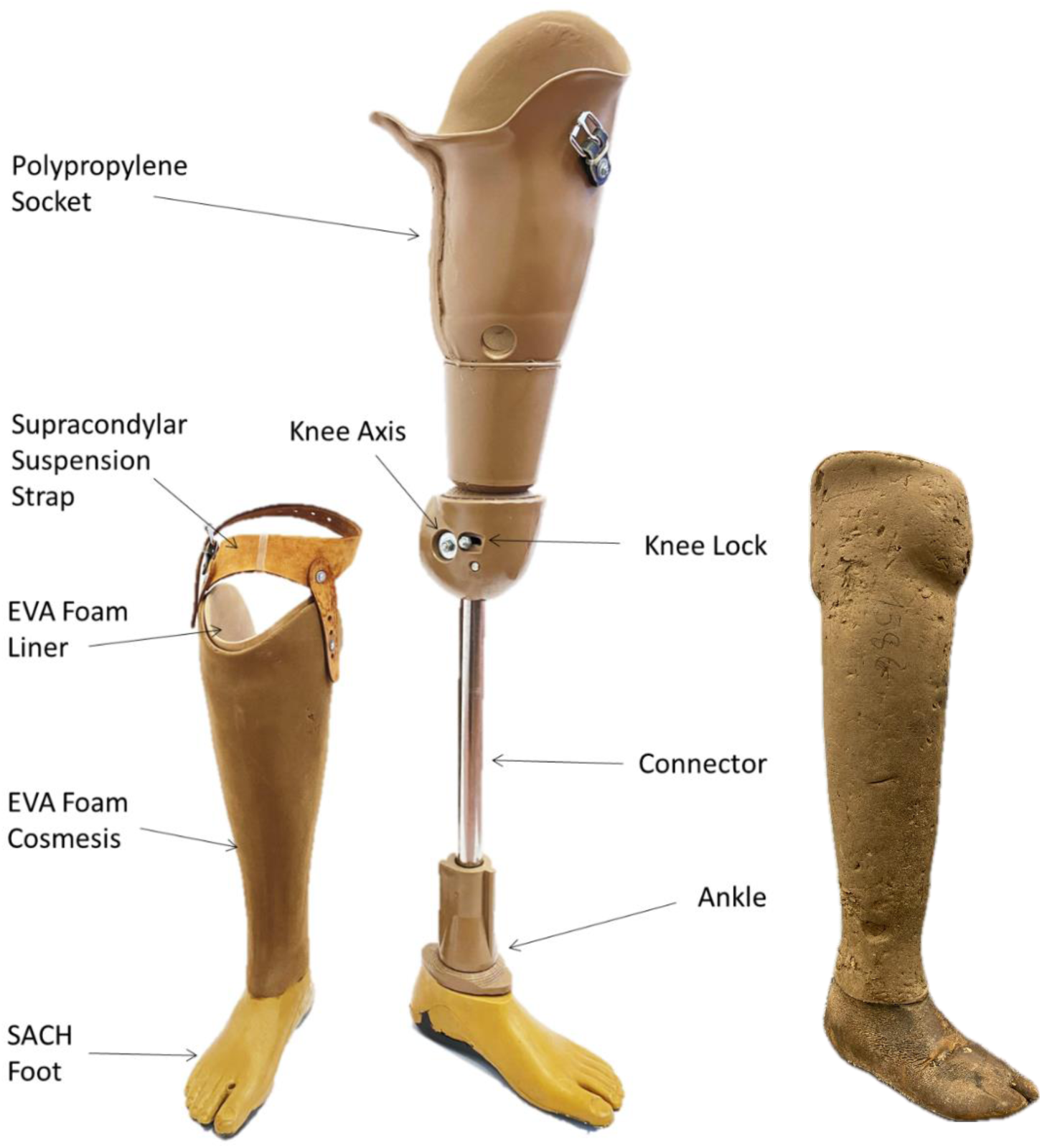
Left: Example polypropylene technology prosthetic limbs produced by Exceed Worldwide Cambodia, according to ICRC Physical Rehabilitation Programme guidelines. Transtibial limb (left) features a cosmetic foam shell which conceals the polypropylene socket, and transfemoral limb (right) is shown without a cosmetic foam shell to reveal structural components. Transfemoral device may also be used with a suspension belt, whose attachment buckle is shown. Right: A transtibial device of Patellar Tendon Bearing Supracondylar Suprapatellar (PTB-SCSP) design, in a worn condition typical of devices returned when clients are issued replacement devices.

The socioeconomic benefits of prosthetic limb provision may be improved with evidence of the durability of the devices’ componentry, and the robustness of prosthetic socket fit which is determined by the socket’s capacity to accommodate fluctuations in residual limb volume and shape changes. However, this evidence is sparse and variable, and there have been relatively few activities to develop large-scale registries describing the care of people with limb absence [8]. This is in stark comparison to other health services that employ medical devices, such as orthopaedic implants, cardiovascular stents and meshes for hernia repair, whose performance data are monitored in the Medical Devices Outcomes Registry (NHS England Digital) and several countries’ National Joint Replacement Registries (NJRRs). In the absence of detailed, comparable and available clinical data, administrative data may provide valuable insights [9]. Collected retrospective studies (Table 1) of prosthetic episodes in high-income settings in the US and UK [10–12] show lower limb devices lasting on average around 2.5 years, with several prosthetist visits per year for socket adjustments or replacement, maintenance and minor repairs. Upper limb devices are reported to last longer before replacement, at 3.5-5yrs, and go longer between visits for maintenance, primarily for repairs [13,14]. Data in low-resource settings are sparse and less current, despite the potentially even greater socioeconomic impact arising from the poor function or failure of a person’s prosthesis. A 1982 study of 25 years of service delivery in India reports the average life of a prosthesis at ‘about 5 years’ [15], and an anecdotal report on prosthetics in ‘Developing Countries’^1^ states that adults need a replacement device typically every 3-5 years [16].

**Table 1:** Exemplar previously published data on frequency of prosthetic device repairs and replacements.

| Study | Context | Years Covered | Findings |
| --- | --- | --- | --- |
| [10,11] | UK, adult, lower limb | 1988-2002 | 2-7 years per device (0.14-0.5 new prostheses per year)<br>1.6-3.4 years per socket (0.29-0.63 new sockets per year)<br>0.23-0.32 major repairs per year<br>1.41-1.70 minor repairs per year |
| [12] | US LEAP, lower limb | 1994-2004 | 2.3 years per device (0.43 new prostheses per year)<br>4-5 prosthetist visits per year |
| [13] | US VA NPPD lower limb | 2000-2010 | 2.5 years between prescriptions<br>1.8 years between repairs |
| [13] | US VA NPPD upper limb | 2000-2010 | 3.6 years between prescriptions<br>4.8 years between repairs |
| [14] | UK, upper limb | 2013-2018 | 0.2 visits per year for a new device (5 years per device)<br>0.9 visits per year for maintenance |
| [15] | India | 1954-1978 | "About 5 years" per device |
| [16] | 'Developing Countries' | 2006 | Anecdotal, 3-5 years per device |

This collected research shows relatively consistent patterns of prosthesis repair and replacement across several settings but indicates the decision to repair or replace is not necessarily driven by device failure. There may be an influence of prescription and funding models, access to services, and in some contexts cultural factors, which influence who is given priority or encouraged to seek health care [15]. It is unknown whether some providers might make different decisions of whether to repair or replace devices depending upon their capacity or skills, and how readily their clients can access the prosthetics clinic. Furthermore, most of the cited studies present the mean number of visits per year or years of life per device. However, those who present more detailed descriptive statistics report high variability within their cohorts, without investigating in depth the underlying reasons.

This study aimed to analyse episodal statistics for a group of prosthetic limb users in Cambodia, to investigate the distribution of the time between device repair and replacement, and to consider variability by studying the influence of a range of user characteristic factors. The research questions were defined as:

- What is the distribution in survivorship of prosthetic devices before repair or replacement?
- Are there inequalities between groups of users in how often an individual gets repairs to their prosthesis?
- Are there differences between groups of users in how often an individual receives a replacement prosthesis?
- Where trends are observed, are there potential health, social, political or technical explanations?

## Methodology

This study undertook an observational retrospective analysis of routinely collected longitudinal data describing people accessing prosthetics services over multiple time points. The provision, repair and replacement of prosthetic devices were investigated. The data referred to clients visiting three clinics run by Non-Governmental Organisation (NGO) Exceed Worldwide, in Phnom Penh (PP), Kampong Som (KS) and Kampong Chhnang (KC), Cambodia, between 1992 and 2019 [17]. Approval was granted by national (230&311NECHR) and institutional ethics review boards (ERGO45577&51898) to analyse episodal statistics extracted from Exceed’s digital clinical records. All records were collected in a standardised manner and stored in the ‘PMS-5’ database (ICRC, Geneva, Switzerland). Each line of data described a single clinical contact such as an assessment, a prosthetic device provision, repair, or replacement. An individual client could have multiple contacts for provision of prosthetic devices, and an individual device could have multiple repairs.

The study used two different groups from the administrative data. Firstly, all records were assessed (from 1992) to understand the services provided over time. Secondly, only individuals who had used any service in the 7 years^2^ prior to 31^st^ December, 2019, were included. This group is classified as ‘active’ users. Factors explored for inequalities were year of birth, gender, occupation or community role, type of prosthesis (as a proxy for limb absence level), year of prosthetic device receipt, reason/cause of limb absence/amputation, and where relevant, the type of prosthesis repair and reason for replacement. Information about an individual on these dimensions were not routinely collected at every visit, so the first record for an individual that did record an answer was used, except for occupation/role which was likely to change, so the most-recently-recorded answer was used. Missing data was grouped into its own categories within each factor, where appropriate, to ensure the sample size was not reduced.

Data analysis was performed in Stata (v17, StataCorp, Texas, USA). As most data were skewed in their distribution, descriptive statistics used the median and interdecile range (IDR, 10^th^-90^th^ percentiles). A Kaplan-Meier survivorship analysis was conducted to describe the distribution of time until repair or replacement of devices. The median time between replacement of prosthetic devices, and the median time between repairs of the same device, was calculated across different groups. Kaplan-Meier analysis does not accommodate time-dependent variables, or control for covariates. Therefore, selected frequencies of repair per group were plotted over time. Differences between Kaplan-Meier estimates of median and quartile survivorship for subgroups were assessed for significance at the 5% level by assessing overlap between the 95% confidence intervals.

To assess the most important factors relating to replacement or repair of devices, the Prentice, Williams and Peterson [18] recurrent event Cox model was used to estimate hazard ratios for repair and replacement separately. This approach is a conditional risk set model, accounting for correlations within the same individual or device for replacements or repairs, allowing the associations with other factors to be examined. Results indicate factors that are significantly related to an increased hazard of a replacement or repair. Finally, trends were reviewed for potential health, demographic and service delivery interpretations.

## Results

### Exploratory Data Analysis

After removing duplicate appointments linked to multiple appointments on the same day for people with bilateral limb absence, the dataset described 51,785 clinical contacts for 6,986 individual clients, of whom 2,894 were classified as active (Appendix Table 4). 14,822 full prosthetic devices were supplied, with an average of 3 (IDR 1–9) devices per person (Table 2). Women were supplied with more devices than men, and there were no clear differences in the number of devices supplied according to their year of birth. People with limb absence for congenital reasons had more prostheses (median 5, IDR 2–14), and people with upper limb amputations received fewer devices (transhumeral, TH: median 1, IDR 1–4 and transradial, TR: median 2, IDR 1–5). Notably, people visiting the Kampong Som clinic received more devices (median 6, IDR 1–13) than clients visiting the other two clinics.

**Table 2:** Median (interdecile range, IDR) number of prosthetic devices delivered to all clients across characteristic groups, and median (IDR) number of repairs per device, with count of deliveries or repairs in each group.

|  |  | *Deliveries* |  | *Repairs* |  |
| --- | --- | --- | --- | --- | --- |
|  |  | **Median (IDR)** | **Count** | **Median (IDR)** | **Count** |
| **Gender** | Girls/Women | 4 (1 – 11) | 2201 | 1 (0 – 3) | 2805 |
|  | Boys/Men | 3 (1 – 8) | 12621 | 1 (0 – 4) | 17029 |
| **Year of Birth** | Pre 1940 | 2 (1 – 8) | 218 | 1 (0 – 2) | 205 |
|  | 1940-1959 | 3 (1 – 8) | 3556 | 1 (0 – 3) | 4456 |
|  | 1960-1969 | 3 (1 – 9) | 3882 | 1 (0 – 4) | 9416 |
|  | 1970-1979 | 3 (1 – 8) | 2253 | 1 (0 – 3) | 3002 |
|  | 1980-1989 | 3 (1 – 9) | 1052 | 1 (0 – 4) | 1758 |
|  | 1990 - | 4 (1 – 11) | 861 | 1 (0 – 3) | 997 |
| **Cause of Amputation** | Congenital | 5 (2 – 14) | 811 | 1 (0 – 3) | 909 |
|  | Traffic Accident | 3 (1 – 8) | 943 | 1 (0 – 5) | 1633 |
|  | Weapon Injury | 3 (1 – 9) | 11752 | 1 (0 – 3) | 15490 |
|  | Animal Bite | 5 (1 – 11) | 213 | 0 (0 – 3) | 251 |
|  | Illness | 2 (1 – 8) | 478 | 1 (0 – 4) | 687 |
|  | Accident at Work | 3 (1 – 8) | 323 | 1 (0 – 4) | 466 |
|  | Other | 2 (1 – 6.5) | 238 | 1 (0 – 3) | 276 |
|  | Missing | 1 (1 – 5) | 64 | 1 (0 – 4) | 122 |
| **Type of Prosthesis** | Partial Foot | 3 (1 – 8) | 306 | 1 (0 – 3) | 374 |
|  | Transtibial (all) | 3 (1 – 9) | 11647 | 1 (0 – 3) | 14996 |
|  | Knee Disarticulation | 4 (1 – 8) | 180 | 1 (0 – 3) | 198 |
|  | Transfemoral | 3 (1 – 8) | 2309 | 1 (0 – 4) | 3836 |
|  | Transradial | 2 (1 – 5) | 212 | 1 (0 – 3) | 215 |
|  | Transhumeral | 1 (1 – 4) | 45 | 1 (0 – 7) | 115 |
|  | Other | 1 (1 – 5) | 105 | 1 (0 – 2) | 95 |
| **Clinic** | Phnom Penh | 3 (1 – 7) | 6510 | 1 (0 – 4) | 11718 |
|  | Kampong Chhnang | 4 (1 – 9) | 4039 | 1 (0 – 4) | 6074 |
|  | Kampong Som | 6 (1 – 13) | 4273 | 0 (0 – 1) | 2042 |
| **Overall** |  | 3 (1 – 9) | 14822 | 1 (0 – 4) | 19834 |

The event records also contained 22,205 repairs (i.e. individual components repaired or replaced) on 19,834 different occasions (several repairs were to multiple different components at the same time). This represented a median of 1 repair visit per device which was the same for almost all demographic and health variables, with IDRs from 0–3 or 4. Exceptions included many fewer repair visits for devices supplied by the Kampong Som clinic (median 0, IDR 0–1) compared to the other two, and devices with more moving parts (transfemoral, with a knee joint, and transhumeral, with an elbow joint) had a larger range in number of repairs (IDRs 0–4 and 0–7, respectively) than the rest.

The time series data (Figure 2 left) indicate a sharp rise in deliveries from 1992 to a peak of 710 deliveries in 1999, a stabilisation in the 2000s at around 500-600 deliveries per year, and a decline in 2018 and 2019. Similar trends are seen for the frequency of repairs over time (Figure 2 right), with approximately a 2–3-year time lag. Since approximately 2006 the three clinics have delivered a similar number of devices, but a greater proportion of the repairs have been undertaken at the Phnom Penh clinic, and a lower proportion at the Kampong Som clinic. Overall, KS delivered 29% of the devices including 34% of the replacements but only 11% of the repairs, compared to PP’s 44% of deliveries, 34% of replacements and 52% of repairs. Finally, a sharp drop in repairs at the Phnom Penh clinic is observed in 2019.

**Figure 2:**
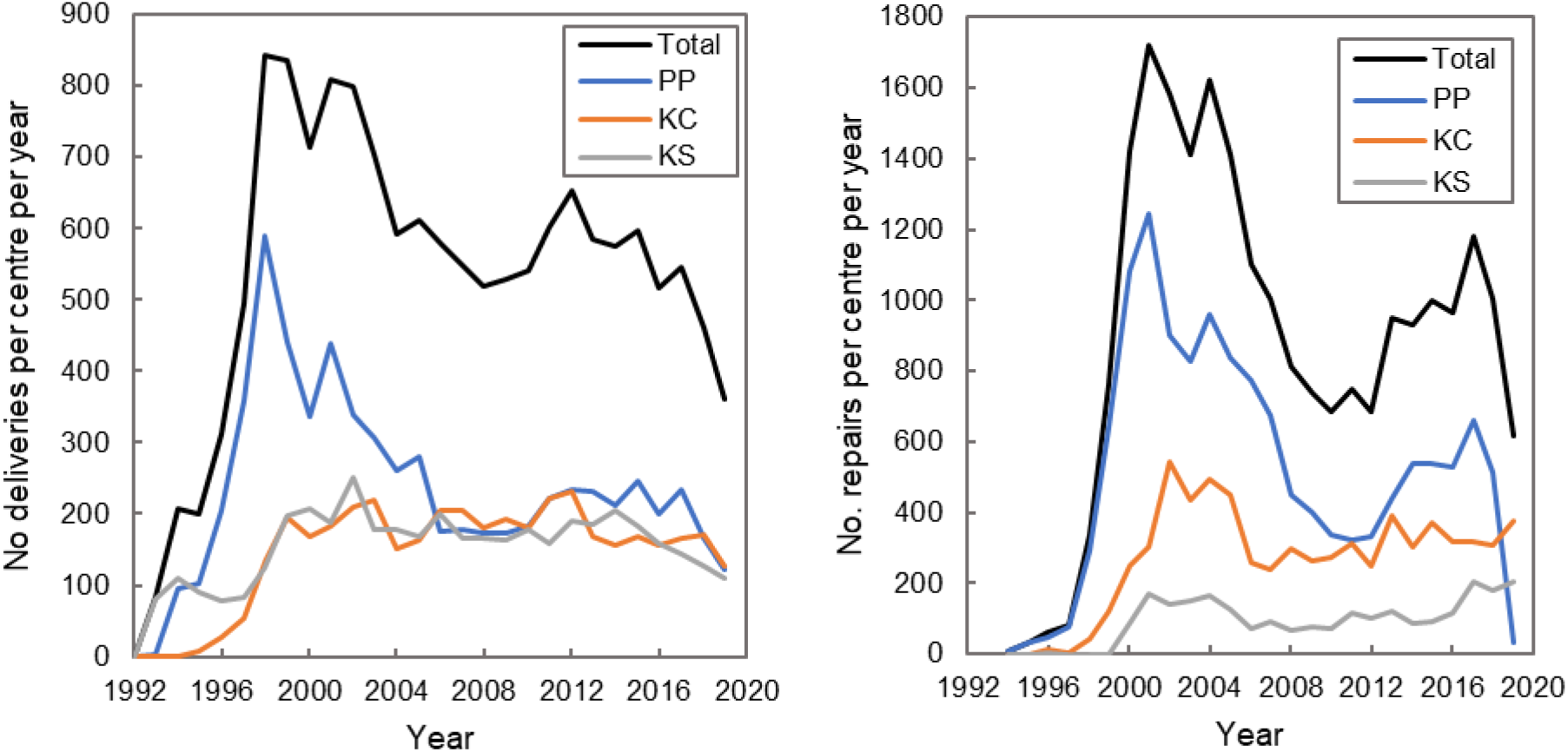
Time series graphs of the number of deliveries (left) and repairs (right) per year, showing the number for each of the three Exceed Worldwide centres and the total. PP denotes Phnom Penh, KC Kampong Chhnang and KS Kampong Som.

### Types of repair and reasons for whole device replacement

From most to least prevalent, repairs included replacement of straps and adjustment of foot alignment, repairs and adjustments to sockets, replacement cosmeses and foot bolts, and replacements to knee, hand or elbow components or their constituent parts (Table 3 top). The initial trend in repairs over time was dominated by strap repairs (Figure 3). In addition, 12,210 socks were provided to manage residual limb volume fluctuation or loss during repairs (although this is likely to be an underestimate of the number as more will have been distributed when a new device is delivered, but were not recorded on the system). Only 99 of the 19,843 different repairs were for socks only, without other repairs occurring at the same time. This indicates that socks are most frequently provided at the same time as other repair events, to replace worn socks or delay the need for socket or limb replacement due to residual limb volume loss, and were therefore not considered as repairs to the prosthetic device and were excluded from the analysis.

**Figure 3:**
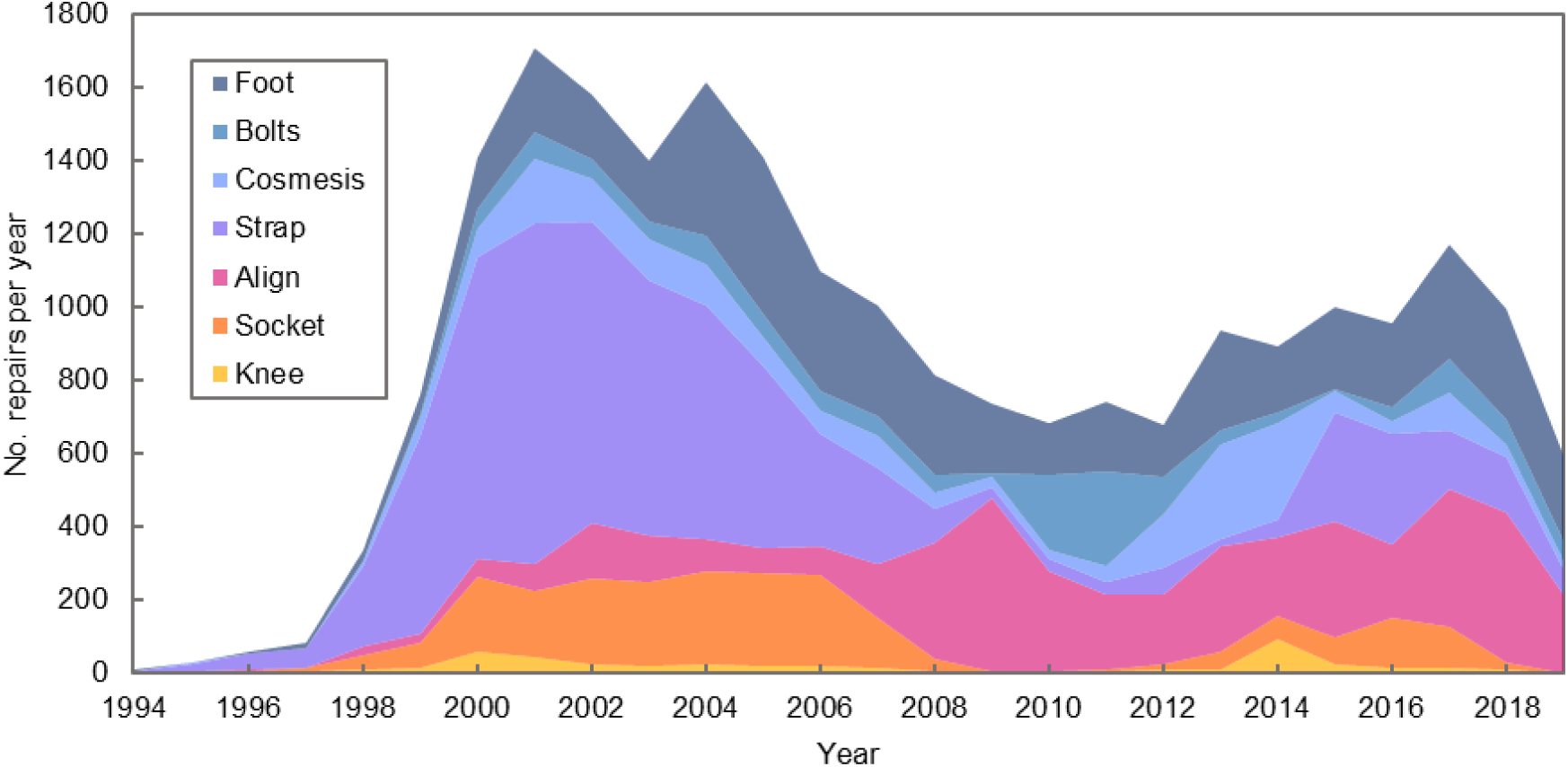
Stacked chart of the number of different repair types provided over time.

**Table 3:** the number of different repair types, and replacement reasons, across the surveyed event records for all clients.

| Centre | Phnom Penh |  | Kampong Chhnang |  | Kampong Som |  | Total |  |
| --- | --- | --- | --- | --- | --- | --- | --- | --- |
| Repair Type | (%) | Count | (%) | Count | (%) | Count | (%) | Count |
| Replacement Strap | 45.9 | 5384 | 23.0 | 1398 | 10.8 | 220 | 35.3 | 7002 |
| Replacement Foot | 17.5 | 2051 | 38.8 | 2359 | 25.0 | 511 | 24.8 | 4921 |
| Alignment adjustment | 12.8 | 1500 | 32.8 | 1992 | 35.1 | 716 | 21.2 | 4208 |
| Socket repair / adjustment | 15.0 | 1759 | 5.9 | 360 | 1.2 | 25 | 10.8 | 2144 |
| Replacement cosmesis | 6.4 | 748 | 12.8 | 780 | 33.6 | 686 | 9.6 | 1914 |
| Replacement foot bolt | 9.6 | 1129 | 1.8 | 112 | 8.0 | 164 | 7.1 | 1405 |
| Replacement knee or component | 3.0 | 351 | 0.7 | 42 | 1.9 | 39 | 2.2 | 432 |
| Replacement hand or component | 0.6 | 73 | 0.5 | 33 | 2.1 | 43 | 0.8 | 149 |
| Replacement elbow or component | 0.1 | 16 | 0.2 | 11 | 0.1 | 3 | 0.2 | 30 |
| Total | 100 | 11724 | 100 | 6077 | 100 | 2042 | 100 | 19843 |
| Replacement Reason | (%) | Count | (%) | Count | (%) | Count | (%) | Count |
| Volume Change | 32.2 | 2094 | 59.8 | 2414 | 66.9 | 2859 | 49.7 | 7367 |
| Wear and Tear | 15.5 | 1012 | 17.6 | 711 | 1.7 | 74 | 12.1 | 1797 |
| Other / False Info | 16.8 | 1098 | 0.0 | 0 | 0.3 | 14 | 7.5 | 1076 |
| Defined Other* | 0.8 | 49 | 0.7 | 29 | 1.2 | 52 | 0.9 | 130 |
| Primary Patient** | 0.8 | 52 | 0.0 | 0 | 0.0 | 0 | 0.4 | 52 |
| Missing | 3.4 | 224 | 0.7 | 28 | 14.9 | 636 | 6.0 | 888 |
| No replacement recorded | 30.4 | 1981 | 21.2 | 857 | 14.9 | 638 | 23.5 | 3476 |
| Total | 100 | 6510 | 100 | 4039 | 100 | 4273 | 100 | 14822 |

\* Defined Other includes Pain, Stolen/Lost, Wrong Alignment and Accident
\*\* This evidences a coding error, as this should not have been used as a reason for a replacement.

**Table 4:** Raw characteristics of people accessing Exceed Worldwide services for prosthetic assessment, prosthetic device provision, repair, and replacement. Percentages include missing data. There are small differences between these two groups, with active clients more likely to be women, seen in Kampong Chhnang clinic, and younger.

|  |  | All Clients | N=6986 | Active Clients | N=2894 |
| --- | --- | --- | --- | --- | --- |
|  |  | (%) | Count | (%) | Count |
| **Gender** | Women | 12.6 | 879 | 15.0 | 434 |
|  | Men | 87.4 | 6107 | 85.0 | 2460 |
| **Clinic** | Phnom Penh | 47.9 | 3346 | 41.5 | 1200 |
|  | Kampong Chhnang | 21.9 | 1528 | 29.5 | 855 |
|  | Kampong Som | 30.2 | 2112 | 29.0 | 839 |
| **Year of Birth (Age at end 2019, years)** | Before 1940 (>79) | 3.1 | 215 | 1.5 | 42 |
|  | 1940-1959 (60-79) | 26.6 | 1861 | 25.2 | 729 |
|  | 1960-1969 (50-59) | 43.4 | 3031 | 40.4 | 1170 |
|  | 1970-1979 (40-49) | 15.4 | 1075 | 13.3 | 385 |
|  | 1980-1989 (30-39) | 6.4 | 446 | 9.7 | 280 |
|  | 1990 and later (≤29) | 5.1 | 358 | 10.0 | 288 |
| **Age at first consultation (years)** | 0-19 | 1.4 | 97 | 2.4 | 69 |
|  | 20-29 | 5.0 | 352 | 6.7 | 195 |
|  | 30-39 | 20.1 | 1405 | 17.8 | 516 |
|  | 40-49 | 35.6 | 2488 | 30.2 | 873 |
|  | 50-59 | 21.4 | 1492 | 21.7 | 629 |
|  | 60+ | 16.5 | 1152 | 21.2 | 612 |
| **Reason for limb absence / Cause of Amputation** | Congenital | 3.4 | 235 | 4.6 | 133 |
|  | Road Traffic Accident | 6.7 | 466 | 11.1 | 320 |
|  | Weapon Injury | 75.3 | 5262 | 64.6 | 1868 |
|  | Animal Bite | 1.0 | 71 | 1.4 | 40 |
|  | Illness* | 5.8 | 408 | 8.1 | 233 |
|  | Accident at Work | 2.6 | 181 | 3.3 | 94 |
|  | Other | 2.7 | 190 | 4.4 | 190 |
|  | Missing | 2.5 | 173 | 2.7 | 78 |
| **Role or Profession (earliest reported)** | Labourer/ Mobile Labourer | 53.8 | 3759 | 58.4 | 1691 |
|  | Farmer | 4.3 | 302 | 9.3 | 268 |
|  | Sedentary Worker/Person with few act | 10.2 | 713 | 9.6 | 278 |
|  | Soldier | 9.5 | 661 | 7.1 | 206 |
|  | Child/Student | 2.2 | 154 | 4.6 | 132 |
|  | Other | 0.8 | 53 | 1.7 | 49 |
|  | Unknown/Missing | 19.2 | 1344 | 9.3 | 268 |
| **Type of prosthesis supplied, as a proxy for level of limb absence or amputation** | Partial Foot | 2.5 | 72 | 1.5 | 42 |
|  | Transtibial** | 61.4 | 4287 | 63.9 | 1850 |
|  | Knee Disarticulation | 1.0 | 72 | 1.5 | 42 |
|  | Transfemoral | 20.5 | 1430 | 18.5 | 535 |
|  | Transradial | 4.8 | 337 | 3.6 | 104 |
|  | Transhumeral | 1.5 | 105 | 1.6 | 45 |
|  | Other | 1.7 | 116 | 1.7 | 50 |
|  | No Prosthesis Supplied | 6.7 | 465 | 6.8 | 196 |

\* Illness for all clients includes diabetes (20.8%), disease (41.9%), gangrene (3.9%), infection (31.6%). Illness for active clients includes diabetes (30.9%), disease (45.9%), gangrene (1.3%), infection (20.2%).
\*\* Transtibial includes PTB (patella tendon bearing), PTB SC (supracondylar) and PTBSCSP (supracondylar, suprapatellar).

Of the 14,822 full devices that were delivered, 11,346 had a recorded replacement (Table 3 bottom). The most common reasons for device replacement were volume change and wear and tear. A small number of devices were replaced due to theft/loss, pain and poor alignment.

### Device Survivorship Analysis

Considering active clients, 14,951 device repairs and 9,032 device replacements were analysed. Survivorship before repair and replacement was illustrated using Kaplan-Meier graphs, which show the estimated probability that devices were surviving unrepaired or unreplaced over time^3^. A quarter of devices had been repaired at 105 days, 50% had been repaired at 237 days and 75% had been repaired at 481 days (Figure 4 left). For an end point of replacement (Figure 4 right), 75% of devices survived to 384 days, 50% of devices survived to 727 days and 25% of devices survived to 1299 days.

**Figure 4:**
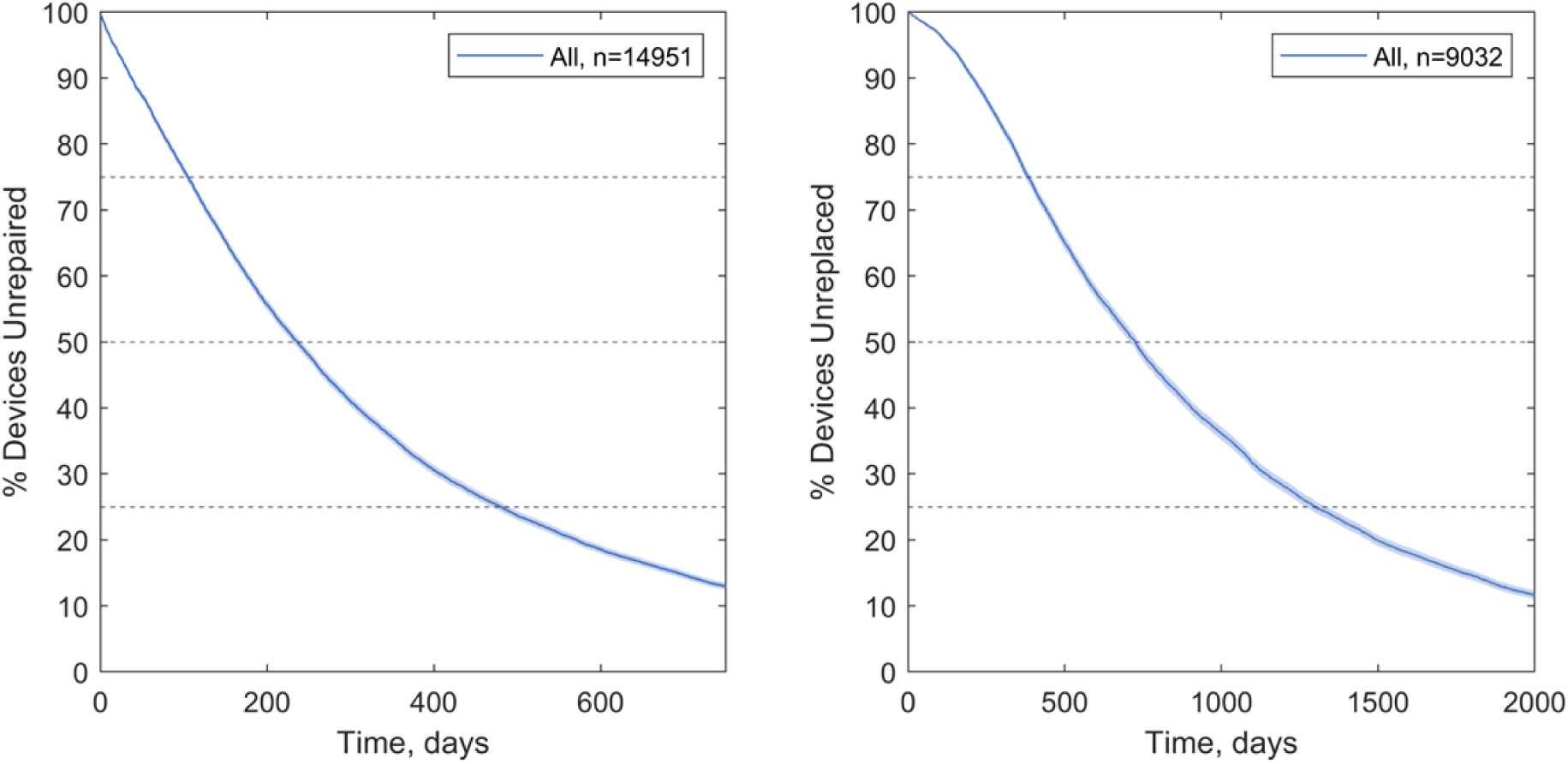
Kaplan-Meier estimates of time to repair (left) and replacement (right) of prosthetic devices, for all active clients. Shaded zones indicate 95% C.I. and dashed lines enable the 25^th^ percentile, median and 75^th^ percentile times to repair or replacement to be identified.

A summary of the time until 50% of devices were repaired and replaced is given in Figure 5 for a variety of client demographic, health, and service delivery dimensions. Kaplan-Meier graphs illustrating noteworthy differences in time to repair and replacement for these subgroups are shown in Figure 7 to Figure 9. Where these dimensions showed no trends, or where survivorship trends were described adequately by the medians in Figure 5, Kaplan-Meier graphs are shown in Appendices (Figure 11 to Figure 18). Finally, to control for confounding between variables, likelihood ratios for repair and replacement were calculated for the same demographic, health, and service delivery dimensions (Figure 6 top and bottom, respectively).

**Figure 5:**
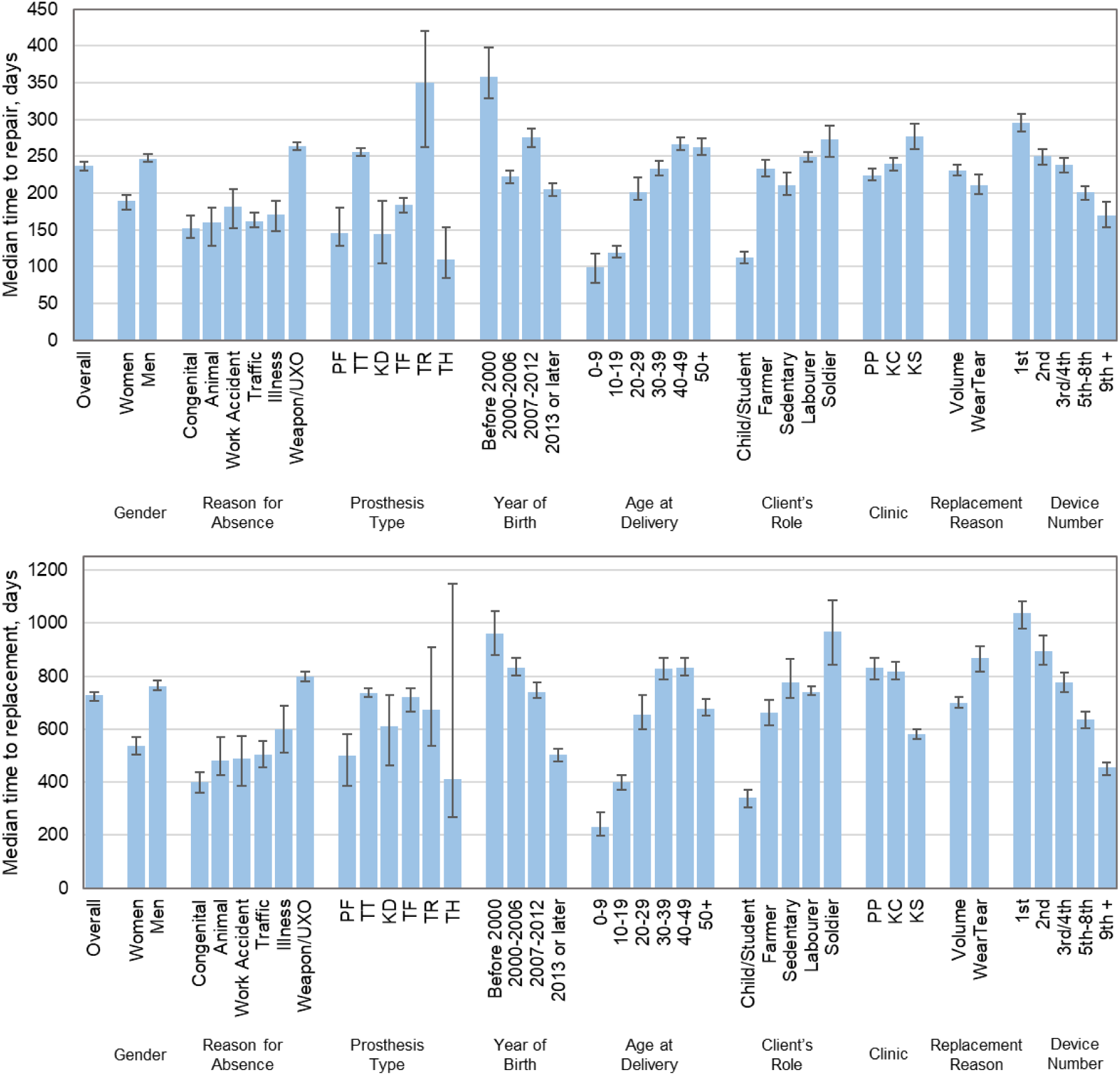
the median number of days between prosthetic device repairs (top) and replacements (bottom), for different population groups, Active clients only. Error bars represent 95% Confidence Interval. Key: Device type: TT = transtibial, TF = transfemoral, PF = partial foot, KD = knee disarticulation, TR = transradial, TH = transhumeral; Clinic: PP = Phnom Penh, KC = Kampong Chhnang, KS = Kampong Som. Note: in repairs (top), Replacement Reason refers to the prosthetic device’s ultimate reason for replacement, and is not an indicator of how the device was repaired.

**Figure 6:**
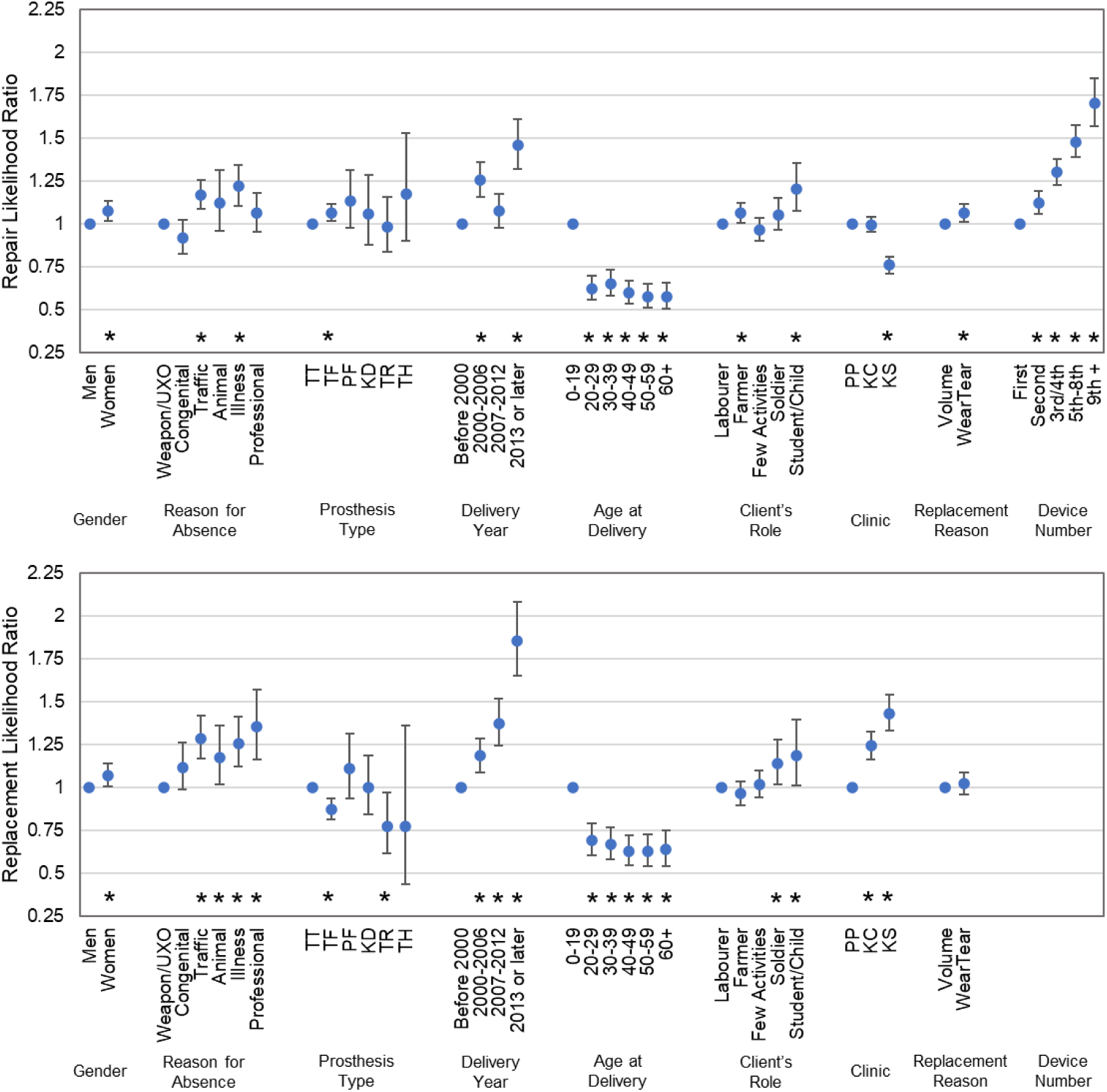
The likelihood ratio of prosthetic device repairs (top) and replacements (bottom), for different population groups. Error bars represent 95% Confidence Interval. Within groups, the reference case is chosen as the most frequent (for e.g. gender, reason for limb absence), or earliest (for e.g. decade of birth or number of device). * denotes statistical significance at p<0.05.

**Figure 7:**
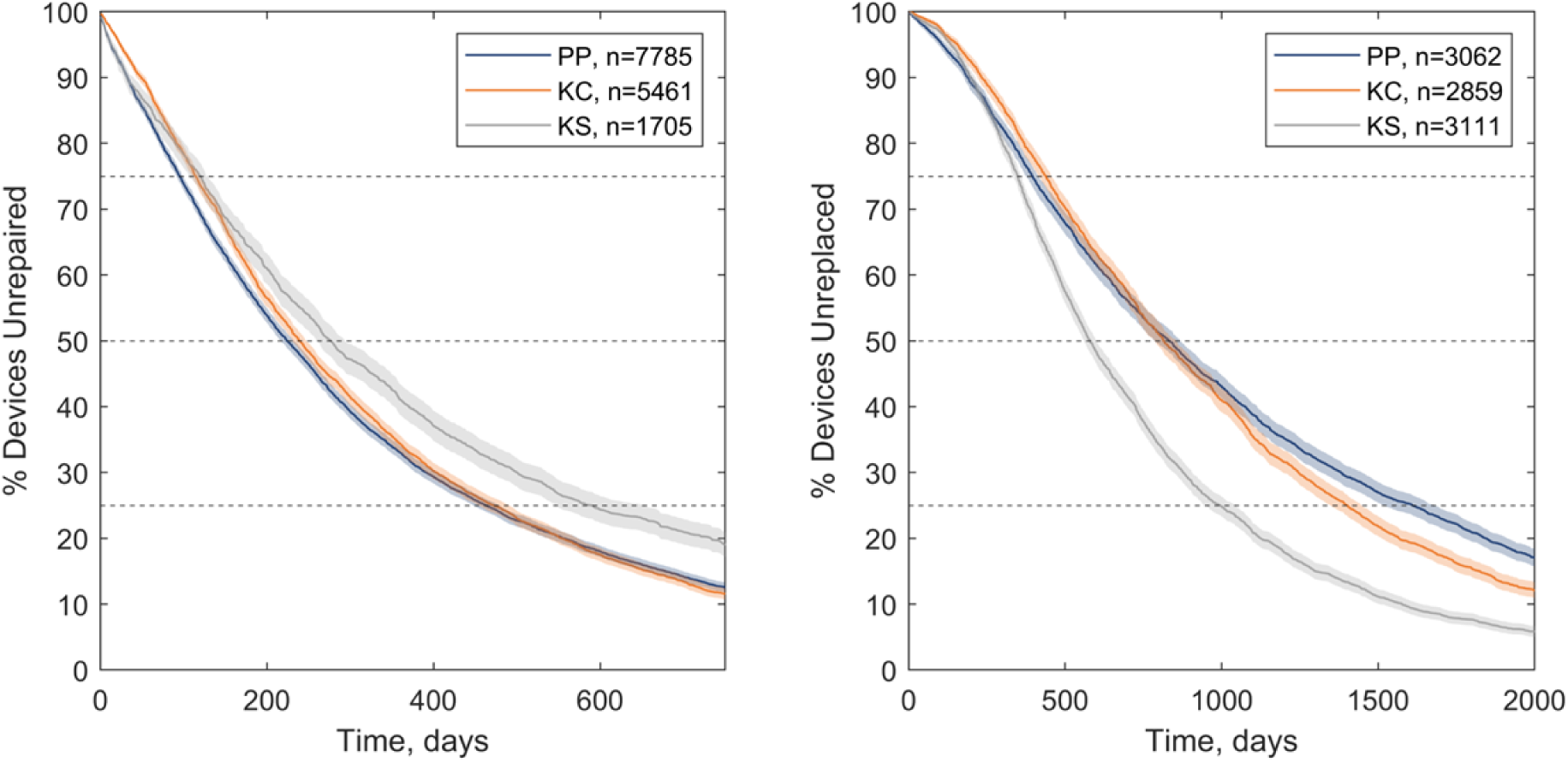
Kaplan-Meier estimates of time to repair (left) and replacement (right) of prosthetic devices, for active clients visiting the three centres run by Exceed Worldwide in Cambodia: PP = Phnom Penh, KC = Kampong Chhnang, KS = Kampong Som. Shaded zones indicate 95% C.I. and dashed lines enable the 25^th^ percentile, median and 75^th^ percentile times to repair or replacement to be compared between groups.

**Figure 8:**
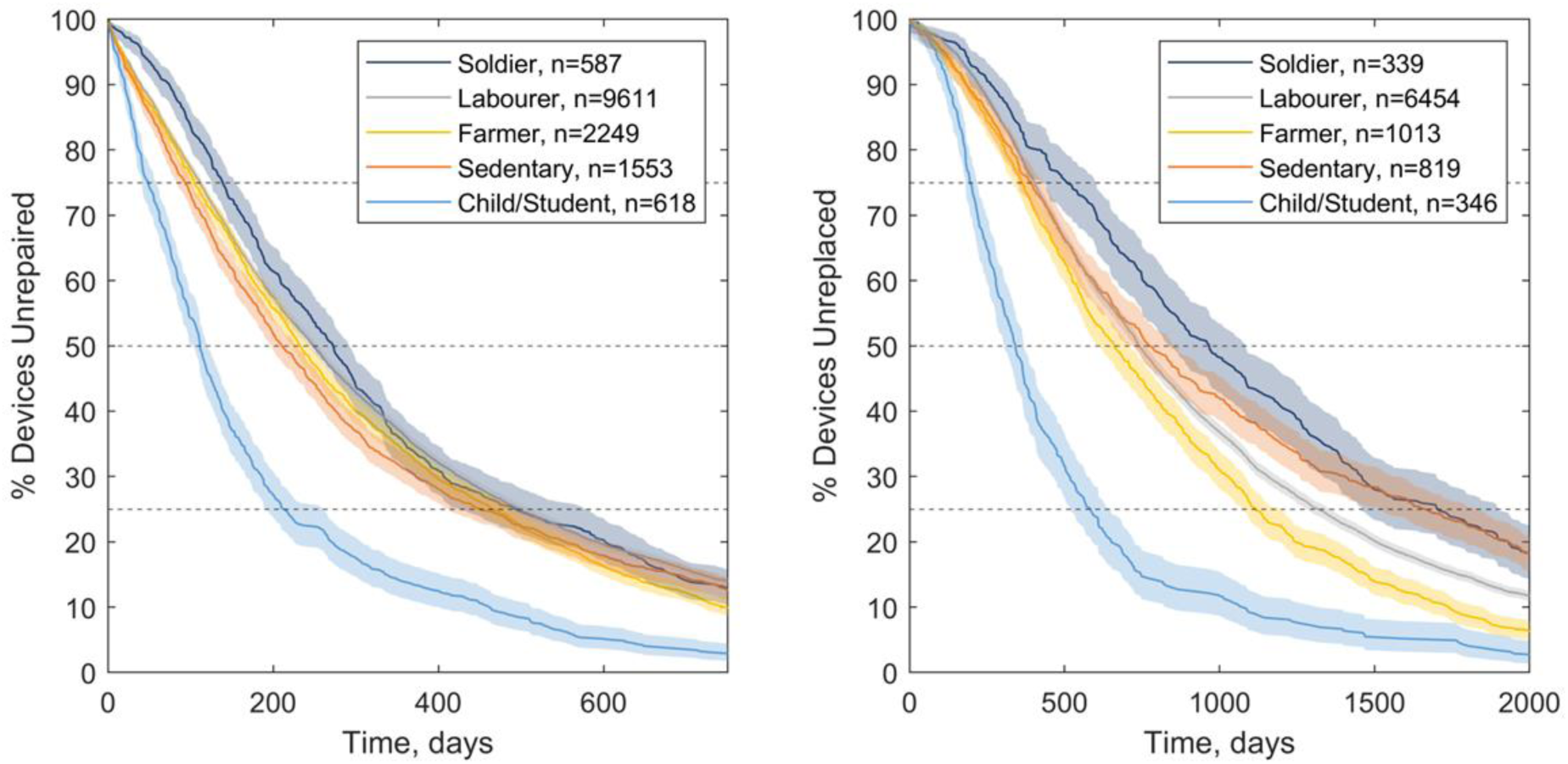
Kaplan-Meier estimates of time to repair (left) and replacement (right) of prosthetic devices, for client occupation or role. Shaded zones indicate 95% C.I. and dashed lines enable the 25^th^ percentile, median and 75^th^ percentile times to repair or replacement to be compared between groups.

**Figure 9:**
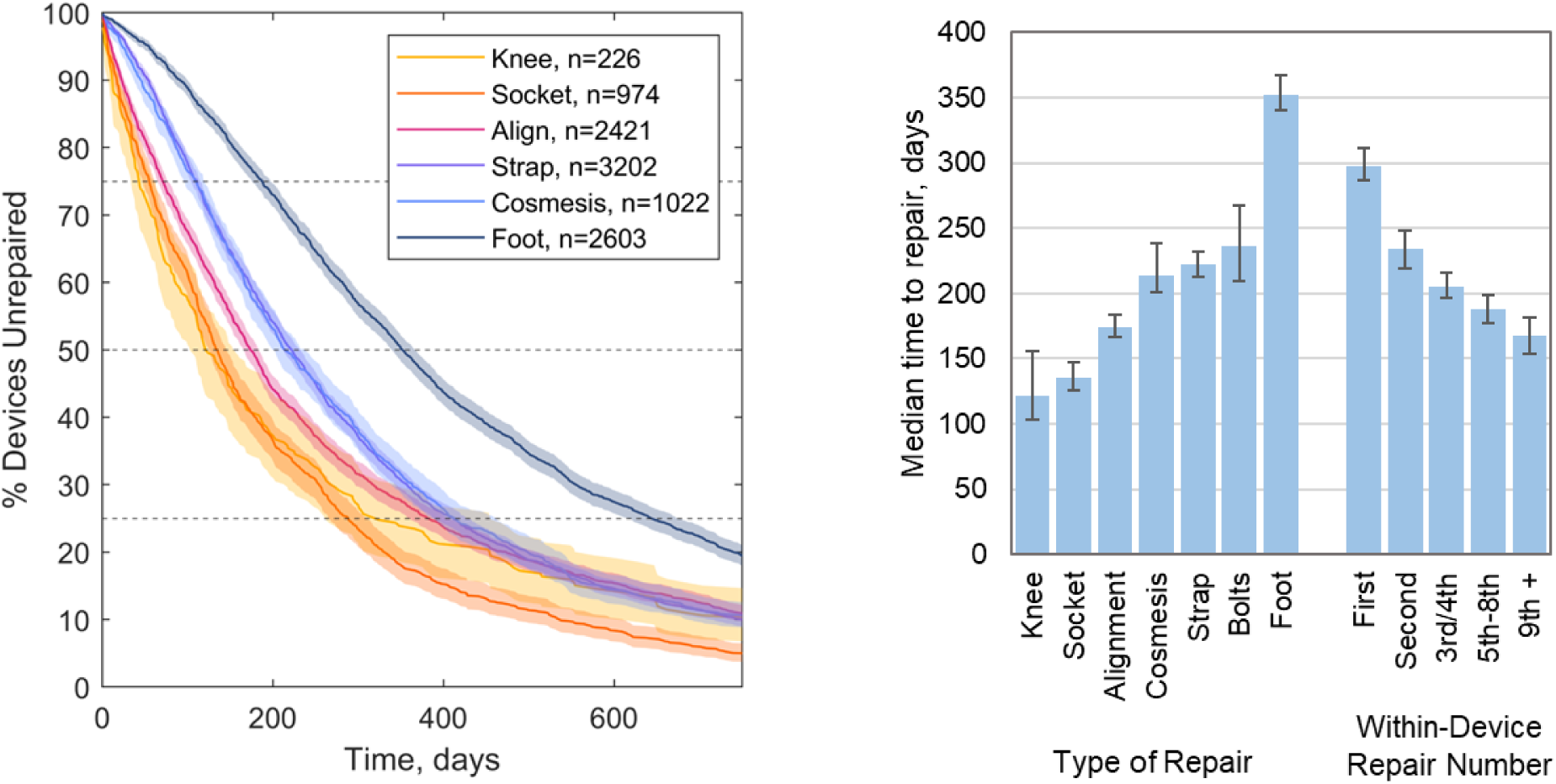
Kaplan-Meier estimates of time to repair of prosthetic devices (top left) and medians (top right), for noteworthy types of repair, and numbers of repair within single devices. Shaded zones indicate 95% C.I. and dashed lines enable the 25^th^ percentile, median and 75^th^ percentile times to repair to be compared between groups.

**Figure 10:**
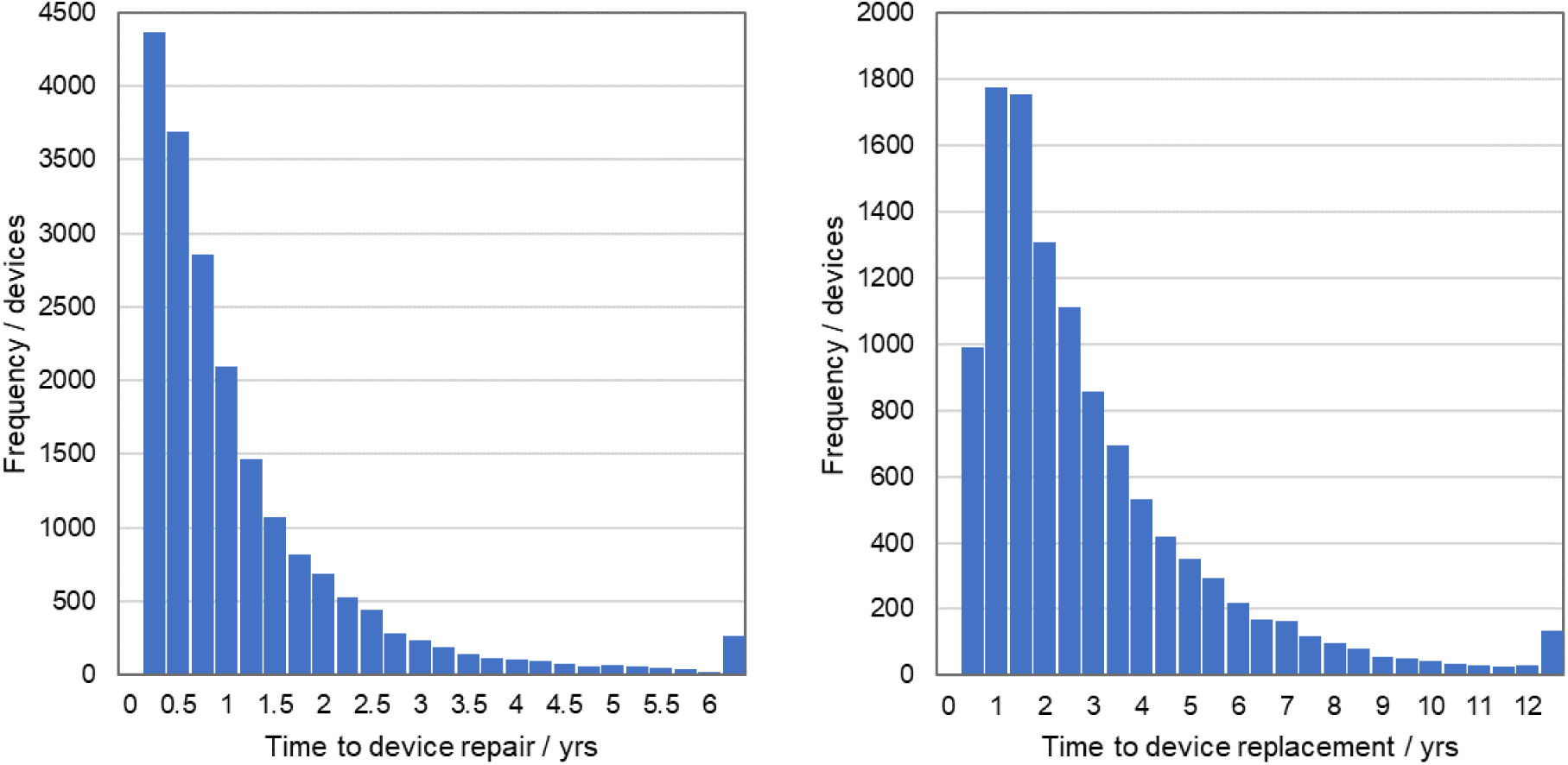
Histograms of time to repair (left) and replacement (right) of prosthetic devices, for all clients

**Figure 11:**
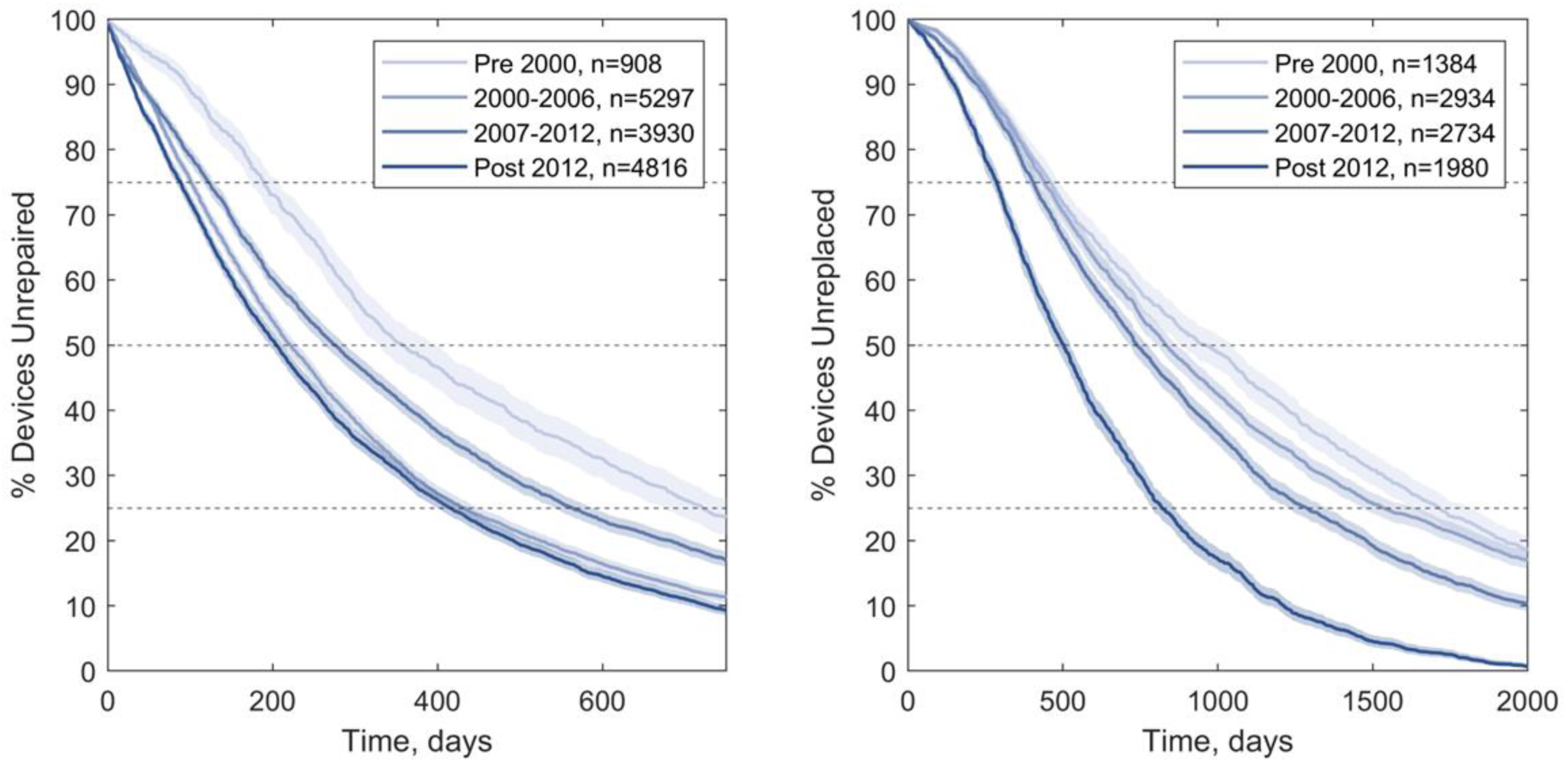
Kaplan-Meier estimates of time to repair (left) and replacement (right) of prosthetic devices, for devices categorised by their year of delivery (top). Shaded zones indicate 95% C.I. and dashed lines enable the 25^th^ percentile, median and 75^th^ percentile times to repair or replacement to be compared between groups.

### Observed Trends and Interpretation: Service Provision Dimensions

Considering service provision dimensions, there is a trend for device repairs and replacements to be provided earlier over time (Delivery Year in Figure 5, Figure 11), and the likelihoods of repair and replacement have also increased (Figure 6). If one assumes that devices are not becoming more unreliable, this may be explained by the prosthetics service becoming more established and better funded, equipped or staffed, and thus having greater capacity. Deviating from this overall trend, a longer time to device repair was observed from 2007-2012 than in the periods either side. This may be explained by the relocation of the largest of Exceed’s three clinics at Phnom Penh between 2008 and 2010, including a period at a temporary site, which could have influenced clinic capacity and client awareness and prioritisation of providing new devices over repairs. This trend is also observed in time series data for the number of repairs and replacements conducted per clinic (Figure 2). A slight drop in device provision is also observed in all three centres in 2018 and 2019, and a marked reduction in repairs at the PP centre in 2019. This might be associated with a reduction in available budget to support clients’ travel costs to access the service, which required Exceed to explore alternative service delivery models. In Phnom Penh this included means testing for cost recovery.

The centres from which clients accessed their prosthetic care was also observed to influence the likelihood and time to device repair or replacement (Figure 6, Figure 7). People served by the Kampong Som centre had a lower likelihood of having their devices repaired than the Phnom Penh and Kampong Chhnang centres, and significantly later, but higher likelihood of replacement, significantly earlier. This may be explained by clients visiting the KS centre from greater distances and more rural areas. Kampong Som is also a coastal province which may expose devices to greater rainfall and a more corrosive environment. As a result, a KS client’s prosthesis may be in a worse condition or beyond repair by the time they can visit the clinic. Prosthetists may also preferentially replace devices instead of making more temporary repair interventions if their client is unable to return frequently or at short notice, due for example to travelling distance or the seasonal workload of subsistence farming [3,19], though analysis of client occupations or roles per clinic indicates relatively few of the KS clients were farmers (Figure 13 left). Differences in the centres’ staff skill makeup and experience in repairing or replacing limbs may also have contributed.

### Observed Trends and Interpretation: Demographic and Health Dimensions

Considering demographic and health dimensions, devices delivered to younger clients had a shorter time to repair and replacement, defined by age at device delivery and decade of birth (Figure 5, Figure 12). The difference was greatest for clients aged 0-9 years, perhaps due to their greater likelihood of limb absence or amputation for congenital reasons and their need for replacement devices due to their growth. While time to repair and replacement was positively associated with decade of birth, the time to repair and replacement was relatively uninfluenced by age at device delivery for adults (over 20 years). Considering gender, women had significantly earlier repairs and replacements than men (Figure 5, Figure 14). This was partially related to their typically younger age than men, but the repair and replacement likelihood was higher for women when these confounding factors were removed (Figure 6). Inspecting the clients’ occupations or roles, those described as ‘child’ or ‘student’ had their devices repaired and replaced significantly earlier than the rest (Figure 8). After accounting for confounding factors (Figure 6), children/students had greater likelihood of repair and replacement than other community roles, and this corresponded with clients who were younger at device delivery, for whom repairs and replacements were significantly more likely than all other age groups.

**Figure 12:**
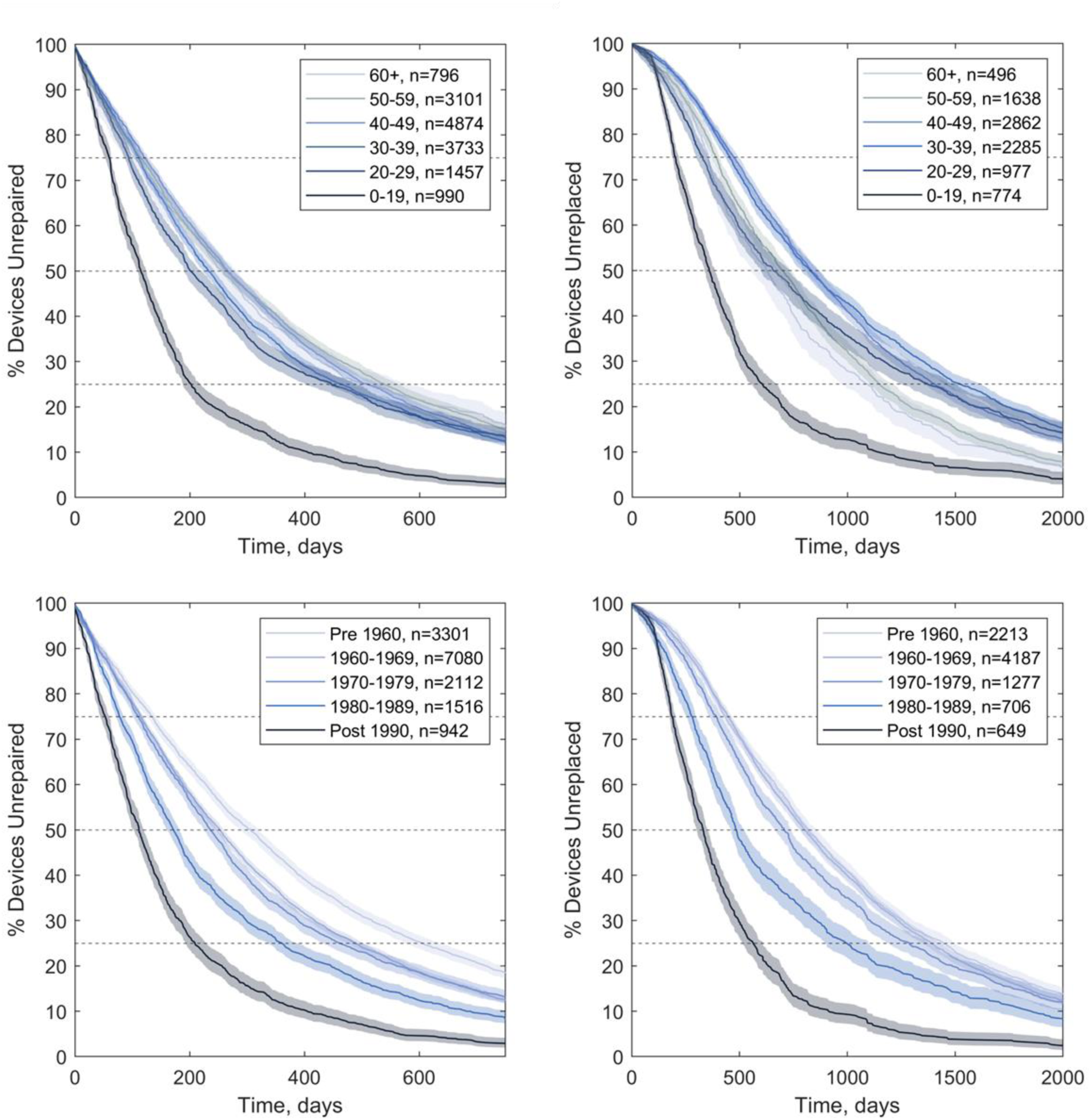
Kaplan-Meier estimates of time to repair (left) and replacement (right) of prosthetic devices, for active clients grouped by their age at device receipt (top) and their birth decades (bottom). Shaded zones indicate 95% C.I. and dashed lines enable the 25^th^ percentile, median and 75^th^ percentile times to repair or replacement to be compared between groups.

**Figure 13:**
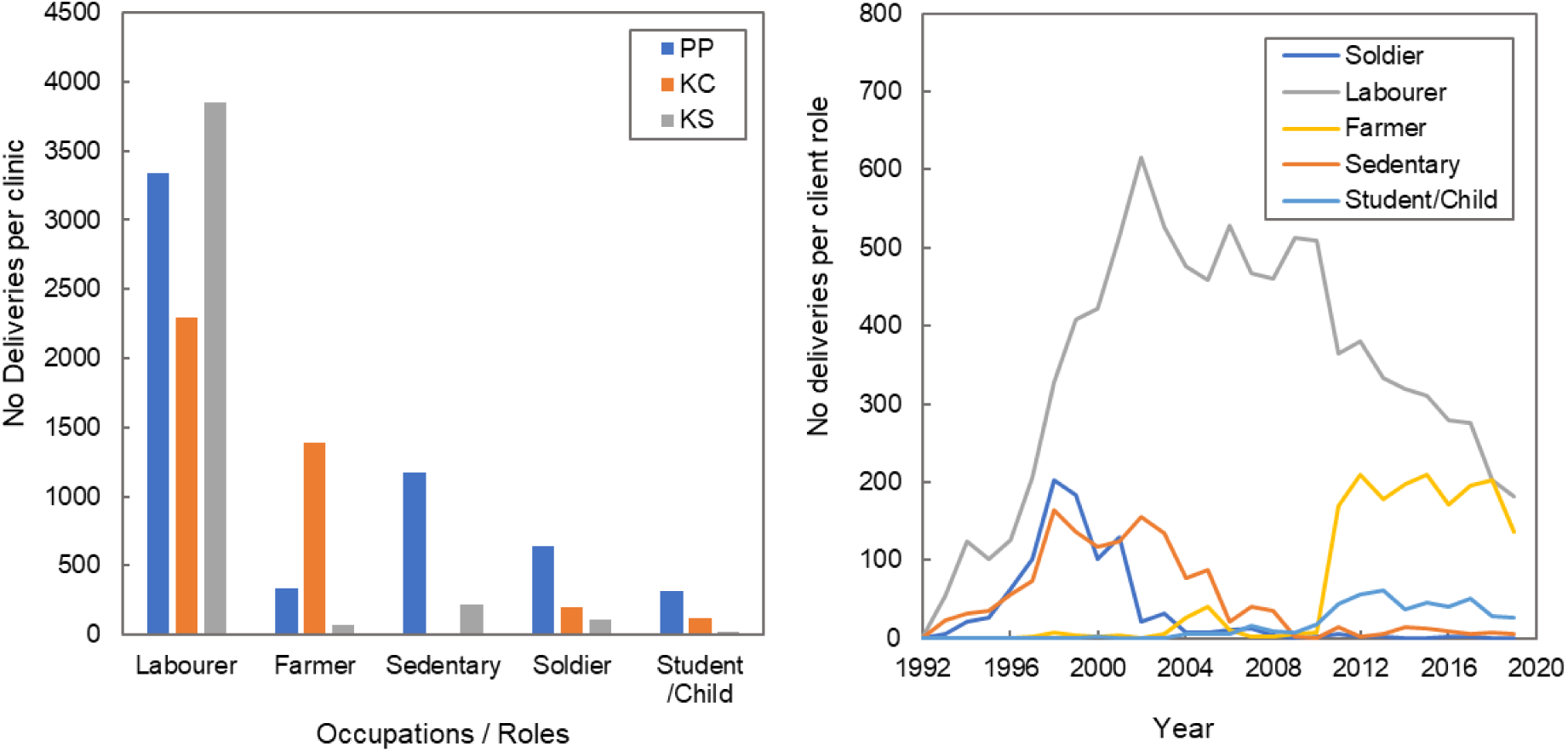
Distribution of client occupations or roles across the three clinics (left) and time series data of deliveries per client occupation or role (right).

**Figure 14:**
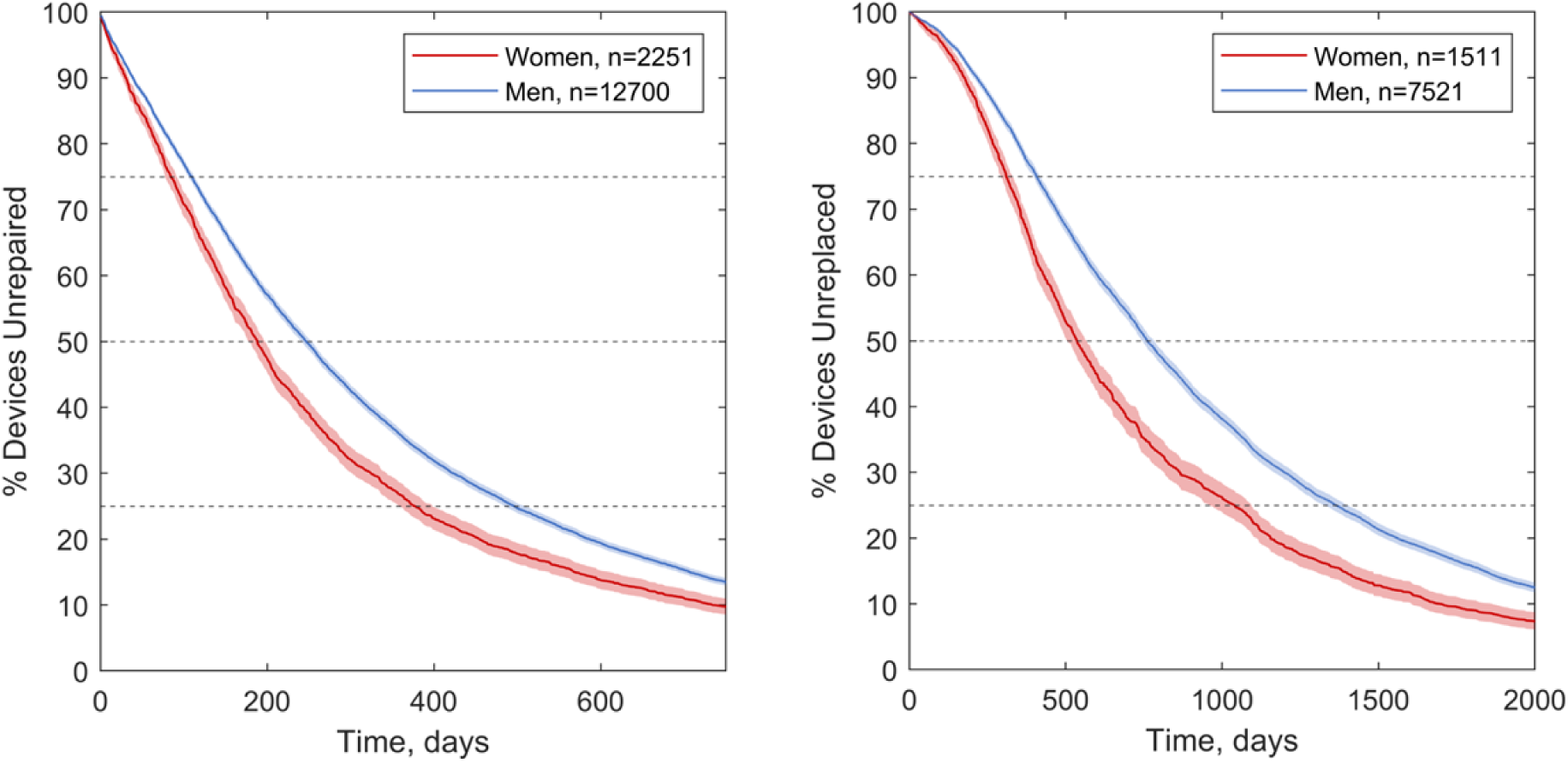
Kaplan-Meier estimates of time to repair (left) and replacement (right) of prosthetic devices, for active clients who were women and men. Shaded zones indicate 95% C.I. and dashed lines enable the 25^th^ percentile, median and 75^th^ percentile times to repair or replacement to be compared between groups.

Similarly, typically older age may explain why people with limb absence due to weapon or unexploded ordinance injury had their devices repaired and replaced significantly later than the other reasons for limb absence, and people using devices due to congenital limb differences had their devices replaced significantly earlier (Figure 15). Controlling for age (Figure 6), these trends in likelihood of repair and replacement for these reasons for limb absence became less strong or disappeared. This indicates that once a person with congenital limb absence reached adulthood, stopped growing and their residual limb stabilised, their devices performed similarly to the rest.

**Figure 15:**
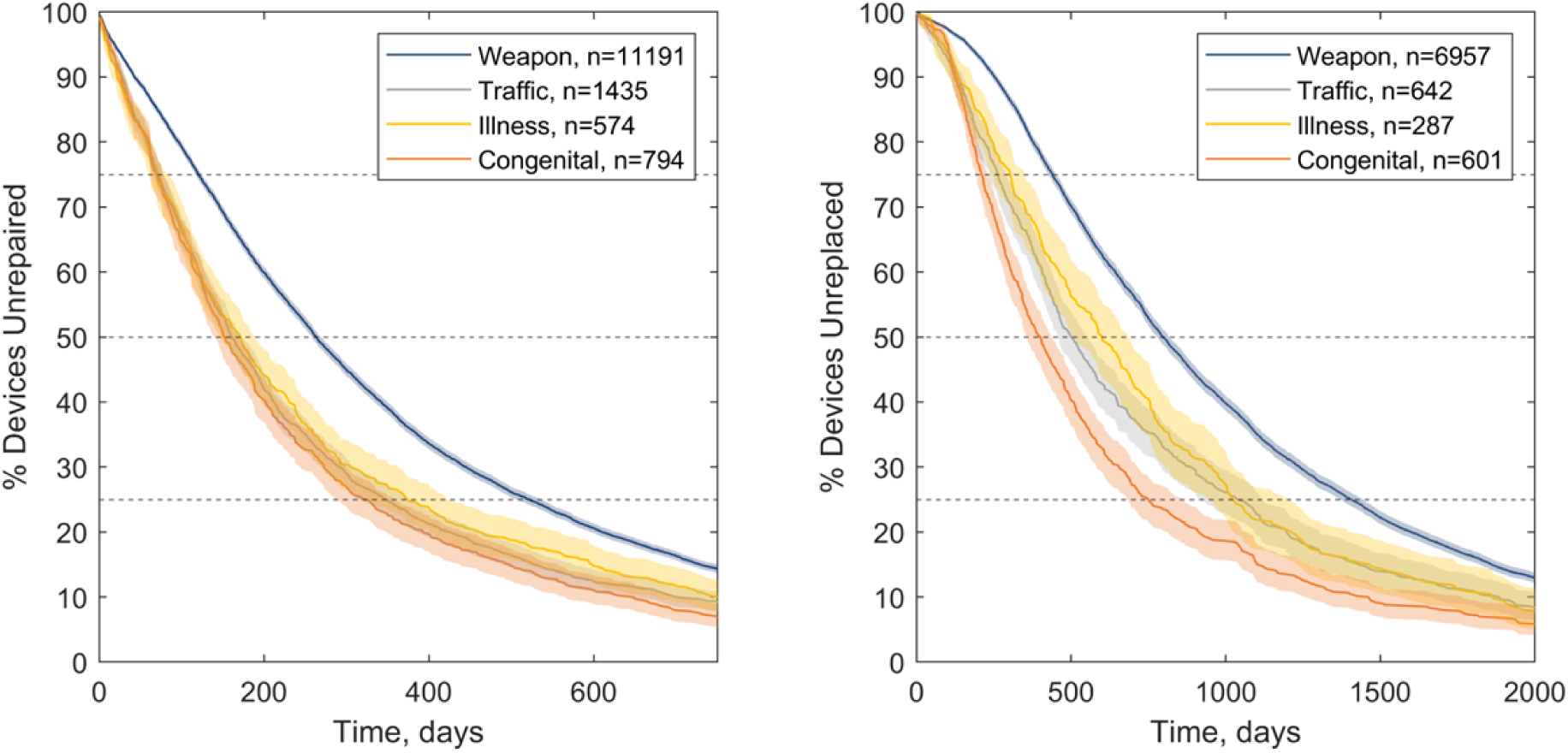
Kaplan-Meier estimates of time to repair (left) and replacement (right) of prosthetic devices, by reason for limb absence in active clients. Shaded zones indicate 95% C.I. and dashed lines enable the 25^th^ percentile, median and 75^th^ percentile times to repair or replacement to be compared between groups.

Considering occupations of adult clients, devices used by farmers and labourers were repaired and replaced earlier than for sedentary people, reaching significance at the 75^th^ percentile survival level (Figure 8), and farmers had a higher likelihood of receiving device repairs (Figure 6). This might be associated with the mechanical forces and environment experienced in their physical work, which is supported by the observation that clients described as sedentary workers or ‘of few activities’ typically received device repairs and replacements later. Devices used by soldiers had significantly longer 25^th^ percentile and median time to device repair replacement, potentially because they are typically the older, more established device users in the study population, and represent the devices delivered earlier in the service’s time period (Figure 13 right). Once correcting for confounding with related variables (number of device, age at delivery and delivery year), soldiers had a similarly increased likelihood of device repair to farmers, and a significantly higher likelihood of device replacement than other labourers (Figure 6).

### Observed Trends and Interpretation: Device Dimensions

Transfemoral devices had significantly greater likelihood of repairs, earlier, than transtibial devices (Figure 5, Figure 16 left), which can be explained by transfemoral devices’ greater technical complexity with more moving parts (i.e. a knee joint and lock) and use of accessories like a suspension strap or belt (Figure 1). There was no significant difference between the replacement survivorship curves for transtibial and transfemoral devices (Figure 16 right), however the Cox model indicated that transfemoral devices did have a significantly lower likelihood of replacement (Figure 6), possibly because their users are generally less active than people with below-knee prostheses. Other device types showed greater variability in survivorship due to smaller sample sizes, but for upper limb devices, transradial limbs were typically repaired later, and more complex transhumeral limbs were repaired earlier. These upper limb devices were less likely to be replaced than lower limbs, reaching significance for the transradial level. This may be associated with the much lower mechanical load placed upon upper- than lower limb devices, and the fact that clients have more choice and opportunity to perform activities of daily living without using their upper limb prosthesis.

**Figure 16:**
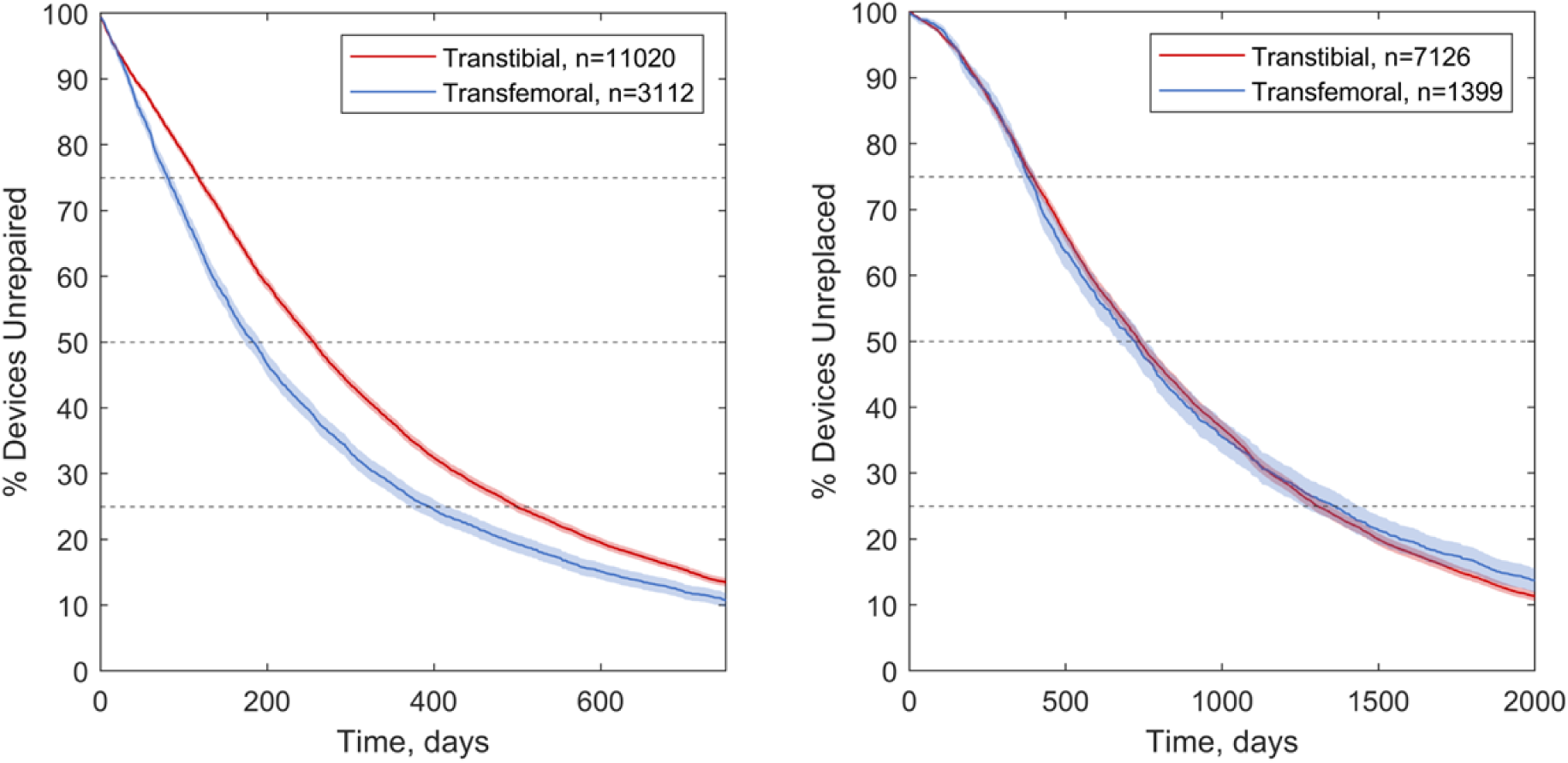
Kaplan-Meier estimates of time to repair (left) and replacement (right) of prosthetic devices, for device type in active clients. Shaded zones indicate 95% C.I. and dashed lines enable the 25^th^ percentile, median and 75^th^ percentile times to repair or replacement to be compared between groups.

When a client had multiple devices, they were typically repaired and replaced progressively earlier (Figure 5, Figure 17). These trends may differ from expectations, where a client might need a large number of devices or repairs by socket adjustments during early prosthetic rehabilitation as their amputation heals and the residual limb volume and shape stabilise, and fewer devices as they become established users. This might be explained by people first accessing prosthetics services a longer time since amputation in low- than high-resourced settings [3,20], at which point their residual limb is more stable, if not yet fully biomechanically adapted to prosthesis use. With limb absence primarily due to trauma from weapon, unexploded ordnance and traffic accidents, clients in this population are more likely to survive for recovery into higher levels of device use than the people with vascular amputations who predominate in high income countries. Thus, they are more likely to return to work and physical activity, exposing their prostheses progressively to heavier wear. Clients who have been accessing prosthetic care for longer may also be more aware of what to ask for and more able to request changes due to discomfort. Further, clients in the present dataset may have previously been receiving care from another physical rehabilitation centre, and what is labelled as their first device is only their first received from an Exceed Worldwide centre [19]. This theory is supported by the Cox model (Figure 6), which revealed a logical trend that multiple devices prescribed to the same person were more likely to experience repairs, indicating progressively higher levels of use as the person becomes a more confident and established limb user.^4^

**Figure 17:**
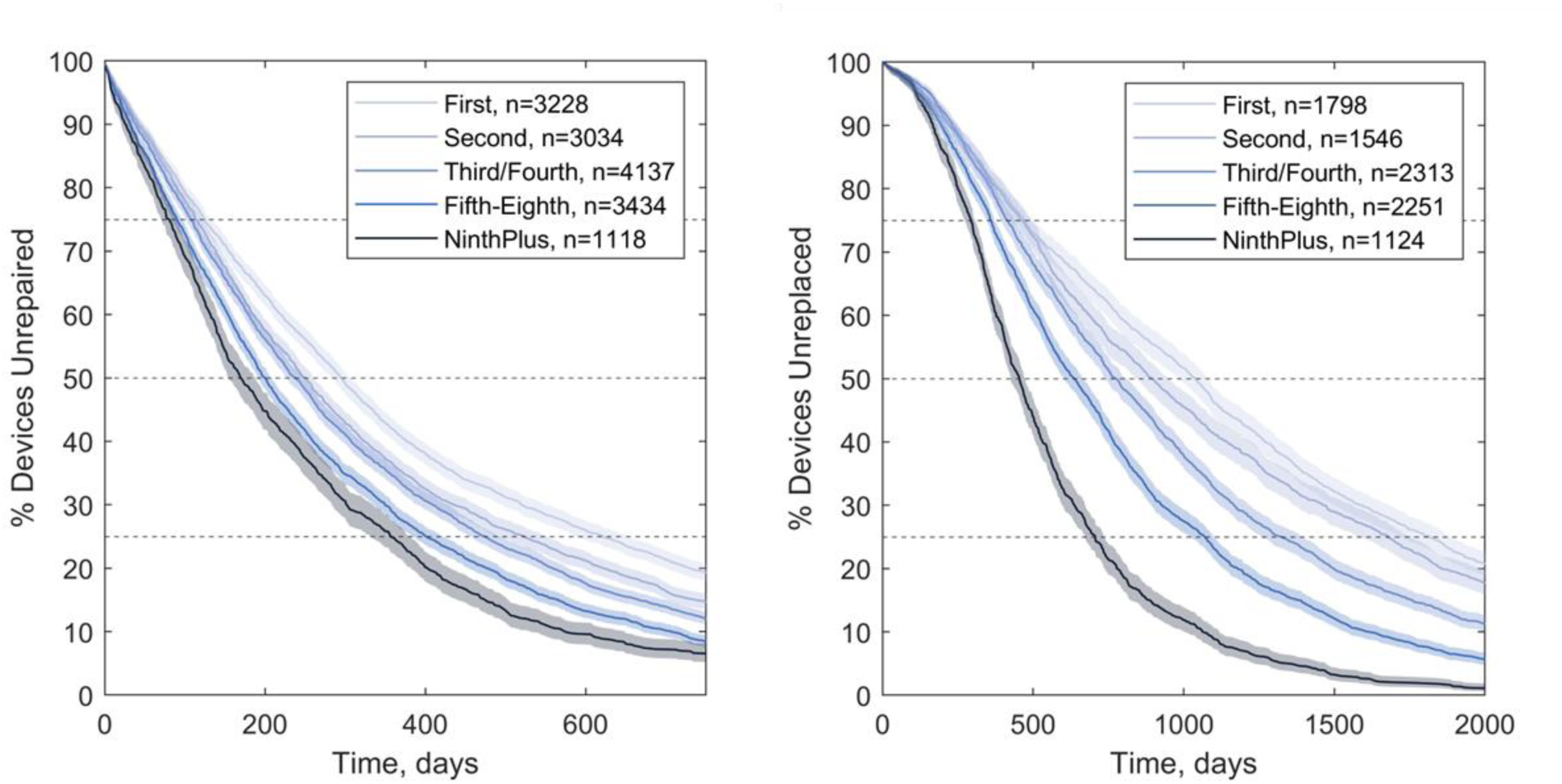
Kaplan-Meier estimates of time to repair (left) and replacement (right) of prosthetic devices, for devices categorised by the client’s device delivery number. Shaded zones indicate 95% C.I. and dashed lines enable the 25^th^ percentile, median and 75^th^ percentile times to repair or replacement to be compared between groups.

Finally, individual device repair and replacement types were considered. The survivorship analysis showed logically-expected trends, that devices which would eventually be replaced due to wear and tear were significantly more likely to be repaired, and sooner than those which would be replaced due to volume change and loss of device fit, whereas device replacement due to volume change occurred significantly sooner than replacement of devices which wore out (Figure 5, Figure 18). Survivorship analysis for different repair types revealing three phases of device maintenance (Figure 9 left). Knee component adjustments and repairs, socket adjustments and device alignments were performed earliest, followed by replacement of suspension straps and cosmeses, whereas device repairs by provision of a replacement foot component occurred latest. The first group might comprise a large proportion of device adjustments in response to initial fitting, with over 15% occurring in the first month following provision, and median times around 6 months when Exceed Worldwide aims to provide their first scheduled follow-up for primary patients. Subsequently, cosmeses and straps are the first device components to be replaced, which is logical as they employ soft materials, are exposed to the environment and are subject to wear and tear. The prosthetic foot survives longest before repair despite its exposure to high levels of mechanical load and wear, because it is critical to the device’s function and therefore is designed to sustain these conditions. The locally made rubber foot has evolved for harsh environments and design decisions to achieve longevity come at the expense of some biomechanical compromises [21]. The relatively early repair by replacing straps and cosmeses may also be due to opportunity, rather than an urgent response to device failure. Where a device had multiple repairs, these occurred progressively sooner (Figure 9 right).

**Figure 18:**
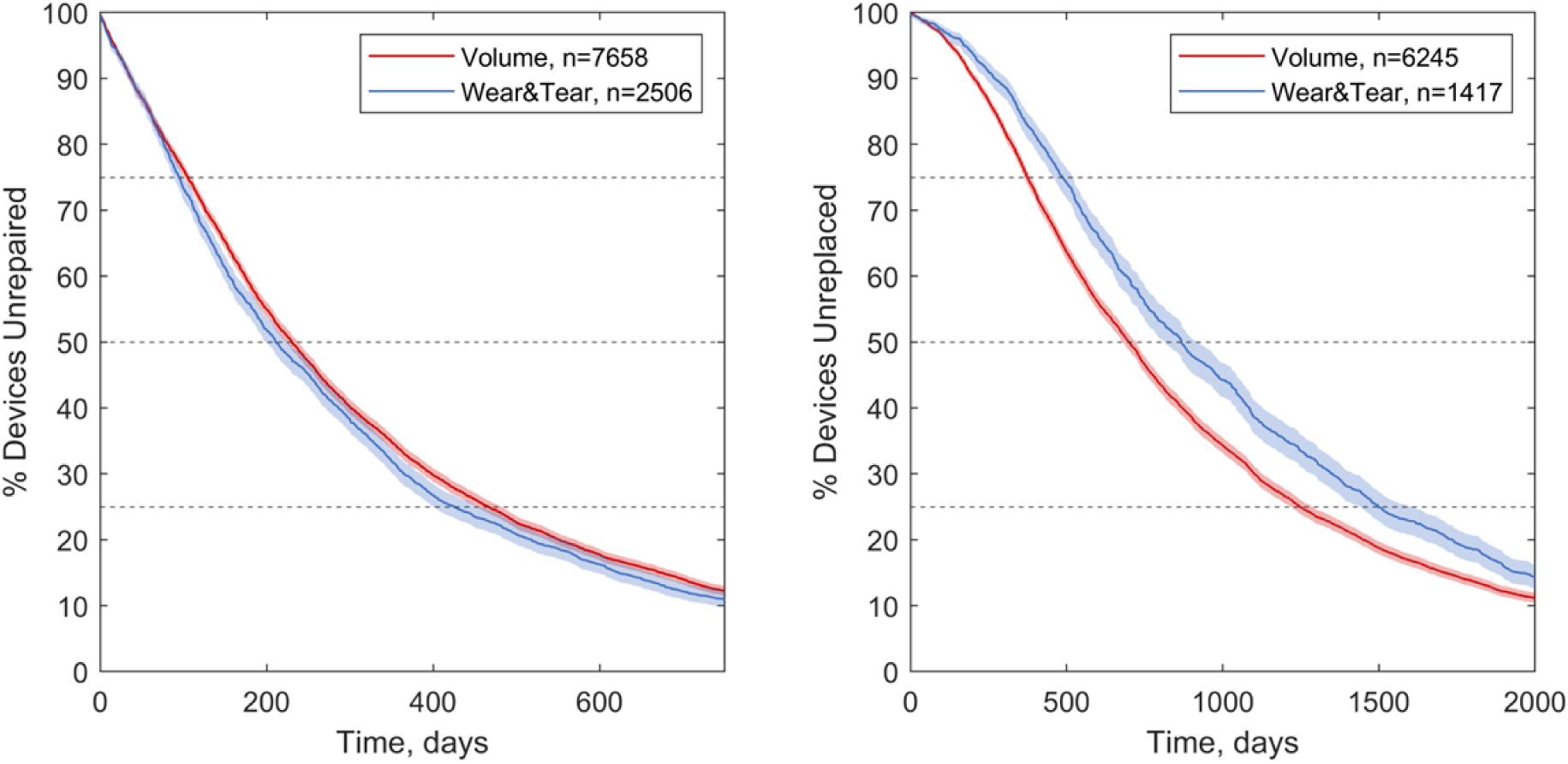
Kaplan-Meier estimates of time to repair (left) and replacement (right) of prosthetic devices, for the two most frequent reasons for replacement. Shaded zones indicate 95% C.I. and dashed lines enable the 25^th^ percentile, median and 75^th^ percentile times to repair or replacement to be compared between groups.

## Discussion

This study analysed the likelihood and distribution of the time until prosthetic devices were repaired and replaced for a group of people with amputations or limb absence in Cambodia, considering the influence of a range of demographic, health, and service delivery factors. Overall, devices were replaced on average after 1.94 years, meaning clients had 0.52 device replacements per year. The average time to device repair was 0.65 years, or 1.53 repairs per year. These are similar to the previously reported data (Table 1) for high income setting services in the UK and US [10–13], and represent considerably more frequent intervention than the historic low resource setting data [15,16], possibly due to Exceed Worldwide’s leverage of international resources and experiences.

The observed higher repair and replacement rates for younger individuals was the same as found in two separate studies, both from a UK context, from 1999, [10] and 2001 [22]. The latter study was also able to identify that more repairs were performed for people with higher body weight, however they saw no correlation between repair rate and gender, cause or level of amputation. Considering differences between devices, a 2008 UK study [11] found that transtibial devices were repaired and replaced more frequently than transfemoral devices, but that the latter had more socket replacements, enabled by device modularity aiding replacement of individual components (categorised in this study as repairs) instead of whole devices. They also observed insignificant trends for prioritising device replacement over repair in younger individuals with transtibial devices, and for more repairs and component changes in people with traumatic than dysvascular amputations. In the present study, the device repair and replacement rate data were highly influenced by the predominant explosive weapon injury cause of amputation and relatively long time since amputation, characteristic of the Cambodian demographic [17,20]. However, when controlling for other variables, repair and replacement likelihoods were significantly higher for people with traffic accident injury and illness (primarily diabetes and infection) than for weapon injury, and these are now the two most common reasons for amputations represented amongst Exceed’s newly presenting clients in Cambodia [17].

Gender also presents an interesting and important dimension. It is established that prosthetic service users are predominantly men in Cambodia and many other low resource settings [17,23] but the implications are unclear. The observed higher likelihood of device repair and replacement for women might imply better access to physical rehabilitation services, or that they need more frequent care. This might be for health reasons or because the predominance of men using prosthetic devices means they have not been specifically designed to suit women’s requirements. Recent studies indicate women are less likely both to have a major amputation and to be successfully fitted with a prosthesis [24], enter prosthetic rehabilitation later [25], and are under- represented in research [26]. The present study cannot distinguish between these possible reasons for gender disparities and this important issue warrants further investigation.

At the component level, a smaller previous study found slightly higher survivorship of the foot, at 100% after 6 months and 80% after 1 year for the ‘HI’ vulcanised rubber Solid Ankle Cushioned Heel (SACH) foot in Cambodia [21], which was listed as the most commonly used foot in the present study (90% of repairs by foot replacement). Finally, the present study agreed with prior findings showing less frequent replacement of upper- than lower-limb devices, as the former experience lower mechanical loads, both in a 1982 low resource setting study [15] and a 2015 analysis of veterans in the USA [13], though the difference in the average time before repair or replacement between upper- and lower limb devices was less marked in this dataset.

Time series data for repairs (Figure 3) indicates several examples of how the service provider has used data collection in the past, responding to observations and improve device reliability by design. For example, the relatively short time before replacement of leather suspension straps was identified by Exceed around 2000, and a decision was then made to promote devices with alternative suspension thereafter (‘supracondylar’ PTB-SC and PTB-SCSP socket designs). The number of strap repairs then declined to a low level around 2009 once these devices had largely been replaced. A small burst of strap repairs was then made around 2015-17, which Exceed attributed to a group of clients with highly demanding occupations requesting strap suspension for added security. Similar benefits of data collection arose in resolving purchased component quality issues. Exceed report anecdotally having observed an increased rate of foot bolt failure around 2010-11 which was attributed to a change in material of the corresponding nut. As observed in the time series data, (Figure 3) following identification of a new supplier, bolt failure incidence returned to a low level. Similarly, perhaps an unexpected result was the short time to repair by knee component as this is a critical, moving and highly-mechanically-loaded part of a transfemoral prosthesis. However, the numbers of knee failures were small, and were attributed to use of plastic in the knee extension block and flexion lock components. Around 2000, these were changed to metallic components and knee failures reduced. Overall, although the ICRC Polypropylene Technologies knee components are functionally simple with their single axis lockable hinge, this study evidences their excellent durability. Prior research indicates a rising incidence and prevalence of transfemoral amputations in Cambodia, so if engineers focus upon new knee prosthesis development for improved function, this dataset sets them an important long-term durability target [17].

## Limitations

The present study is subject to some limitations. Such analysis is subject to inaccuracies arising from human data input and interpretation of fields. There is some missing data, notably in the clients’ role or occupation, and this may have been completed as their current occupation, or that at the point of amputation. Further possible evidence of variations in how different staff and centres interpreted or used the form includes very low reports of wear and tear as a replacement reason from the KS clinic compared to the other two, and greater use of ‘Missing’, ‘Other’ or ‘False Info’ returns at PP and KS than KC (Table 3). Differences in recording events between clinics may explain some of the reasons for their wide variation, and training is required to ensure similar approaches in recording data especially between centres and countries.

Most devices are constructed by standardised techniques using comparable materials and componentry, which has demonstrated positive outcomes based on activity and quality of life in both qualitative [3] and quantitative [27] research. The study population does include a variety of feet; these were all of SACH design but obtained from different domestic manufacture or import suppliers, for which a range of durability has been reported [21,28]. The present dataset recorded the source of the replacement foot, but not that of the foot which was replaced, so a comparative analysis of the survivorship of the different foot types was not possible.

Moreover, the study does not give a definitive measure of the lifespan of prosthetic devices, because a long time to repair or replacement may not always indicate a positive outcome. This is similar to Joint Registries for orthopaedic implants [29] which have a definite end-point of revision surgery, and this may be considered too high a risk especially for patients who are elderly or have comorbidities. In the present study there are several reasons devices may have failed without being repaired or replaced. Some clients may have stopped using their prostheses, died, or taken their nationally-standardised Polypropylene Technology devices for repair or replacement at centres other than the three included in the study [19]. To avoid clients lost to follow-up from skewing the survivorship, devices without a record of replacement were removed from the survivorship part of the study, and only active clients were considered.

### Interpreting the time to device repair and replacement: a ‘Goldilocks’ optimal time window

Care must be taken over interpreting early repair or replacement of a heavily worn device as always representing a negative outcome. Heavy wear may indicate a device is comfortable and functional, and hence is heavily used, which anecdotally prosthetists report as a success. The wear may reflect the individual’s activity level, occupation and environment; a device worn by a more sedentary office worker will not be the same as that of an agricultural worker. A poorly-fitted, uncomfortable device will not exhibit the same degree of wear. Additionally, device condition and repair or replacement are influenced both by the need for and the opportunity to access a prosthetics clinic. A relatively prompt repair or replacement can indicate that an individual has good access to prosthetics care, without necessarily indicating a shortcoming in device design or fabrication quality. The present study can only provide limited evidence to analyse these technical factors, or the barriers preventing physical rehabilitation centre access. The study also captures only repairs conducted in a formal rehabilitation service setting, whereas the importance of more informal and community-based repair is becoming appreciated, especially in remote and lower resourced settings [4].

Similarly, a very long time to repair or replacement might be considered an undesirable outcome if it results from the person not using their device. There may therefore be an optimal range of time for replacement. Data provided by this study, together with conversations with service providers may enable identification of early repairs or replacements which were opportunistic, or were associated with premature device failure or loss of fit, so that the cause can be addressed. This might help identify how long a device ideally lasts, subject to typical use and access to services for repair or replacement, and therefore define when a device has been repaired or replaced ‘too soon’, especially for clients of particular occupations, implying some premature failure.

## Conclusions

The potential value and risks of use of ‘Big Data’ for health improvements in LMICs [30] are now well established. However, the scientific literature reporting of prosthetic limb survivorship before repair or replacement remains sparse and shows high variability. The present study offers a highly detailed analysis of a relatively large and heterogeneous population of individuals within a single country, all receiving devices from the same service-providing NGO, and comparable devices constructed from ICRC-initiated Polypropylene Technologies with SACH feet.

The high variability of survivorship measures is to be expected with great variety in prosthesis user characteristics and, unlike an orthopaedic implant, a device whose continued use is the person’s choice. However, such analysis will enable comparison with other user groups and settings and may enable us to identify groups who are less well-served by current devices or rehabilitation models, and modify physical rehabilitation services to address these inequalities towards Universal Health Coverage. Device survivorship data may also contribute to cost effectiveness analysis of current services, which is of particular importance for reporting back to government and charitable funders [31] and justifying sustained funding to provide continuity of care.

Finally, the present study provides benchmark data against which engineers could measure new technologies. All new healthcare technologies present a risk, no matter how well-intentioned. The potential impacts of this risk are most severe for vulnerable people in LMIC communities [32], and it is our responsibility to ensure that prosthetics technology innovation offers a genuine improvement above established devices.

## Data Availability

All data produced in the present study are available upon reasonable request to the corresponding author's institution

## Acknowledgments

The authors thank Exceed Worldwide for facilitating, and the Exceed Research Network (ERN) for providing critical review. We also thank the University of Southampton’s Institute for Life Sciences / FortisNet interdisciplinary musculoskeletal research network for supporting our preliminary work.

## Ethical Approval and Informed Consent Statements

The secondary data analysis study was conducted according to the guidelines of the Declaration of Helsinki, and was granted approval by Institutional (ERGO45577&51898) and UK national review boards (230&311NECHR), who waived the requirement for individual consent.

## Footnotes

1 Throughout, we use the phrase ‘low resource settings’.

2 7 years was chosen based on the Exceed Country Director’s recommendation, SK

3 On a Kaplan-Meier survivorship graph, a steeper line indicates that devices are being repaired or replaced sooner than a shallower line. Horizontal lines at the 75th percentile, median (50th percentile) and 25th percentile indicate the estimated time to repair or replacement of 25%, 50% and 75% of devices, respectively.

4 The ‘Device Number’ is not included as a variable in the repeated events replacement likelihood analysis due to the choice of the Prentice, Williams and Peterson model, which uses the device number as the strata.

